# Case finding of early pregnancies at risk of preeclampsia using maternal blood leptin/ceramide ratio: multi-omics discovery and validation from a longitudinal study

**DOI:** 10.1101/2020.12.17.20248418

**Authors:** Qianyang Huang, Shiying Hao, Jin You, Xiaoming Yao, Zhen Li, James Schilling, Zhen Li, Sheeno Thyparambil, Wei-li Liao, Xin Zhou, Lihong Mo, Subhashini Ladella, David Fan, John C. Whitin, Harvey J. Cohen, Doff B. McElhinney, Ronald J. Wong, Gary M. Shaw, David K. Stevenson, Karl G. Sylvester, Xuefeng B. Ling

## Abstract

**Objective:** To evaluate whether longitudinal measurements of serological adipokines and sphingolipids can predict preeclampsia early in gestation.

**Design:** Retrospective multi-omics discovery and longitudinal validation.

**Setting:** Maternity units in two US hospitals.

**Methods:** A multi-omics approach integrating genomic and lipidomic discoveries was employed to identify leptin (Lep) and ceramide (Cer) as novel PE early gestational biomarkers. The levels of placental growth factor (PlGF), soluble fms-like tyrosine kinase (sFlt-1), Lep, and Cer in maternal sera were then determined by enzyme-linked immunosorbent (ELISA) and liquid chromatography-tandem mass spectrometric (LC/MS/MS) assays.

**Main outcome measures:** Interval from positive prediction to confirmative diagnosis.

**Results:** Genomic meta-analysis compiled six PE placental cohorts with 78 PE and 95 non-PE control placentas. The Testing Cohort included sera from 7 non-PE and 8 PE women collected at confirmatory diagnosis. The Validation Cohort included sera from 20 non-PE and 20 PE women collected longitudinally through gestation. Our findings revealed a marked elevation of maternal serum Leptin/Ceramide (d18:1/25:0) ratio from early gestation (a median of 23 weeks) when comparing later PE-complicated with uncomplicated pregnancies. The maternal Lep/Cer (d18:1/25:0) ratio significantly outperformed the established sFlt-1/PlGF ratio in predicting PE for sensitivity (85% vs. 40%), positive predictive value (89% vs. 42%), and AUC (0.92 vs. 0.52) from 5 to 25 weeks of gestation.

**Conclusions:** Non-invasive longitudinal assessment by serological evaluation of Lep/Cer (d18:1/25:0) ratio can case find early pregnancies at risk of preeclampsia, outperforming sFlt-1/PlGF ratio test.

**Tweetable abstract:** Non-invasive longitudinal assessment by serological evaluation of Lep and Cer ratio can predict preeclampsia early in gestation.

## INTRODUCTION

Preeclampsia (PE) is a disorder of the placental vasculature, affecting 5% to 8% of all pregnancies worldwide. It still remains a leading cause of maternal and fetal mortality (1), accounting for 42% of all maternal deaths and 15% of preterm deliveries (2,3). It is characterized by diffused endothelial dysfunction, increased peripheral vascular resistance, hypertension, proteinuria, and dysregulated coagulation. The pathogenesis of PE is complex as it progresses from asymptomatic stage in the first trimester to a symptomatic stage late in gestation. Although its etiologies remain largely unknown, mounting evidence has revealed that placental dysfunction is integral to the development of PE (4). Pathophysiological perturbations of placental development cause incomplete remodeling of the uterine spiral arteries and poor invasion of trophoblasts into placental cells, which induces persistent placental oxidative stress and hypoxia, such as PE (5,6). From a clinical perspective, early prediction of preeclampsia (i.e., within the first 16 weeks of gestion) is of critical importance as it would allow for early treatment of high-risk women, which has been proposed to reduce the occurrence of preeclampsia. Gestational interventions such as steroids to accelerate fetal lung maturity (7), magnesium for seizure prophylaxis (8), aspirin treatment, and antihypertensive therapy (9) are effective in reducing both maternal and fetal mortality in populations with high risks of developing PE (10,11).

However, the early prediction of PE remains challenging. Traditional risk factors such as a prior history of PE, first pregnancy, multiple gestation, and obesity have insufficient sensitivity and specificity (less than 60%) for the prediction of PE (12–15). An imbalance of angiogenic and anti-angiogenic factors during pregnancy was found to disrupt the developmental homeostasis of the placenta (16,17). Two placental-derived factors, angiogenic soluble fms-like tyrosine kinase (sFlt-1) and anti-angiogenic placental growth factor (PlGF), were associated with the pathophysiology of PE (18). A multicenter trial demonstrated that the sFlt-1/PlGF ratio in maternal sera significantly differentiates pregnant PE from normal pregnant women after 24 wks’ of gestation (19,20). Later studies discovered that this ratio had limited value in predicting the development of PE when examined during the first or early second trimesters (21,22). Thus, there is an unmet need to identify sensitive and specific markers to predict PE early in gestation.

Previous studies have suggested that PE is a pregnancy complication that is associated with changes of multiple systems and encompasses genetic, proteomic, and metabolic factors (23–25). Recent multi-omics studies identified a number of molecular-level candidates associated with PE (26–33). One of these candidates is leptin (Lep), a secreted adipokine that affects the central regulation of energy homeostasis, neuroendocrine function, and cytoplasmic metabolism (34). Lep can be expressed by both adipose and non-adipose tissues, which, during pregnancy, not only mediates the gestational energy homeostasis (35), but also modulates various physiological events, such as implantation, placentation, and immune adaption, that are essential for fetal development (36). Our previous findings have demonstrated elevations of Lep in early gestation in PE patients (22,24,37,38). Other have also reported that sphingolipid metabolism, particularly via ceramide (Cer), acts downstream to the anorectic actions of central Lep, and played an important role in Lep-induced hypothalamic control of feeding (39–41). Furthermore, our recent findings have also illustrated the regulatory role of Cer as a metabolic messenger for the homeostatic development of normal pregnancy along gestation (42), and placental changes in cytoplasmic amount of Cer in trophoblast cells have been shown to be implicated in the pathogenesis of PE (43–45).

In this study, we employed a multi-omics approach to identify Lep and Cer as potential biomarker candidates for risk of impending PE. This initial omics-based discovery led to the generation of our hypothesis that the gestational profiles of Lep and Cer differ in maternal serum from women without PE compared with those with PE. We further hypothesized that the ratio of Lep and Cer can serve as a serological marker capable of predicting impending PE early in gestation. We therefore characterized the serological profiles of circulating Lep and Cer longitudinally and investigated their potential utility in predicting impending PE early in pregnancy and biological insights.

## MATERIALS and METHODS

### Study Design

The overall sample allocation, hypothesis generation, biomarker discovery, independent validation, and panel construction workflows are illustrated in Fig. 1. Our study was conducted in three phases: (1) the discovery phase, which included both in-silico expression analysis of PE and non-PE placentas and comprehensive literature mining to generated the hypothesis that Lep and Cer might be implicated in PE pathophysiology as potential biomarkers; (2) the testing phase, which measured the levels of Lep and Cer in a case-control cohort of PE and non-PE maternal sera sampled at confirmative diagnosis; (3) the validation phase, which determined the levels of Lep and Cer in an independent longitudinal cohort of PE and non-PE maternal sera sampled at different gestational ages.

**Figure 1.**
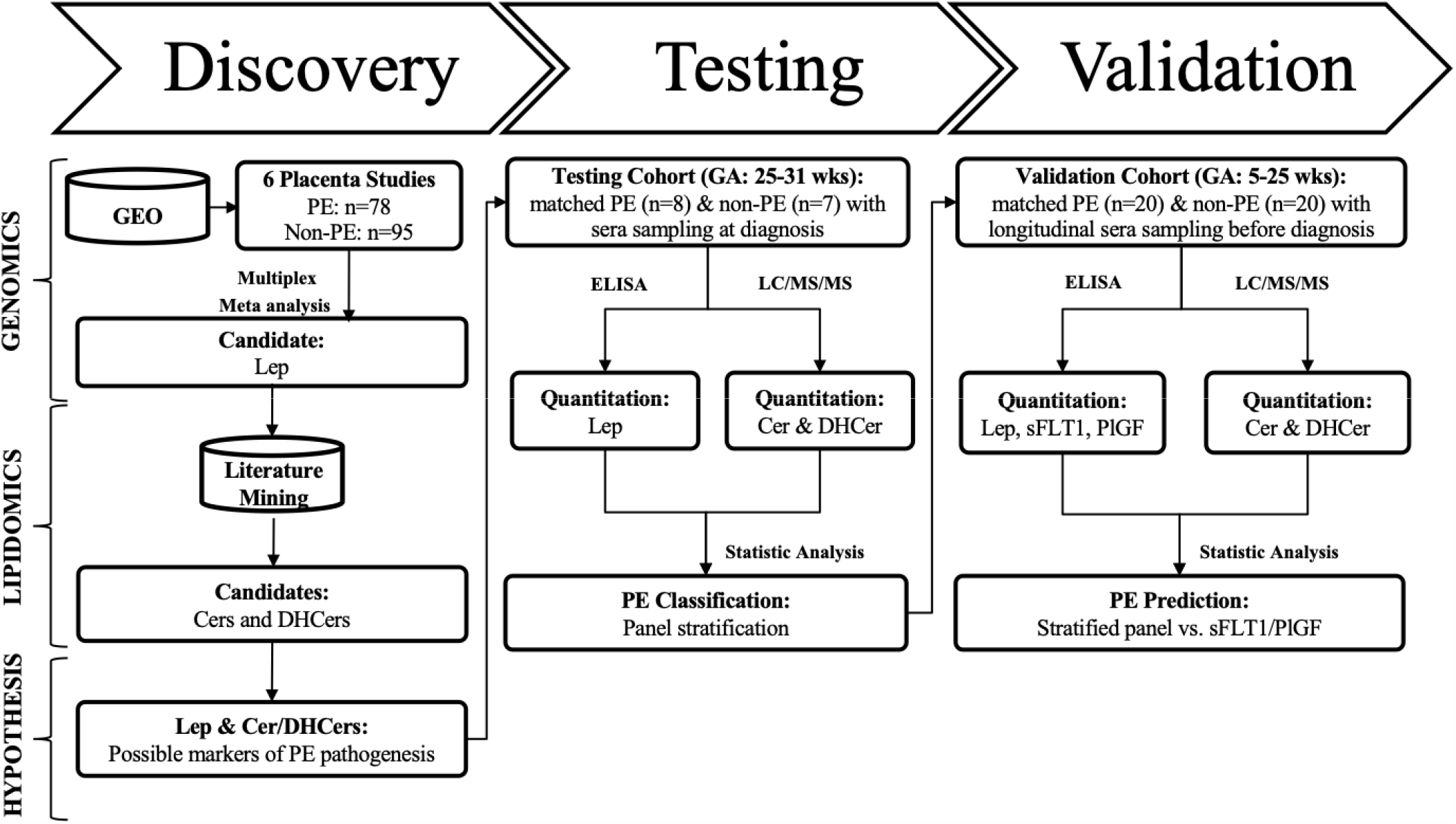
Schematic diagram of the study design.

### Meta-analysis of placental gene expression

Six PE placenta expression studies from Gene Expression Omnibus (GEO) datasets (46–51) were combined and subjected to multiplex analysis with the method as we previously developed (52,53). We calculated the meta-fold change of each gene across all studies. For gene expression measurements, this corresponds to combining fold-changes across studies to identify a meta-fold-change that is an amalgamation of the constituent studies. The meta p-values were obtained by Fisher’s method. Significant genes were selected if they were measured in five or more studies and the meta effect p value was less than 4.5×10^−5^. This effort identified Lep as the leading protein biomarker candidate. We also compared the expression of Lep transcript between non-PE and PE women in individual datasets.

### Study population and blood collection

Two independent cohorts of pregnant women were assembled for this study. The testing cohort included sera from women between 25 and 31 wks’ of gestation with or without PE collected by ProMedDX Inc. (Norton, MA). Each woman had one sample collected. All PE samples were collected at the time of confirmative diagnosis. The validation cohort included sera from women who participated in a longitudinal study sponsored by the March of Dimes Prematurity Center at Stanford University between November 2012 and May 2016. Each woman had multiple sera collected from 5 to 29 wks’ of gestation prior to PE diagnosis. Study approval was obtained from ProMedDX Inc. and the Institutional Review Board at Stanford University. Written informed consent was obtained from all participants.

The diagnosis of PE was made according to the American College of Obstetricians and Gynecologists criteria (54) as follows: a persistent systolic blood pressure ≥140 mmHg, or a diastolic blood pressure ≥ 90 mmHg after 20 wks’ of gestation in a woman having a previous normal blood pressure in conjunction with one or more of the following: new-onset proteinuria, new-onset thrombocytopenia, impaired liver function, renal insufficiency, pulmonary edema, or visual or cerebral disturbances in the absence of proteinuria. Early-onset PE was defined as PE that develops before 34 wks’ of gestation, whereas the late-onset PE develops at or after 34 wks’ of gestation.

### Enzyme-linked immunosorbent assay (ELISA)

Serum concentrations of Lep, sFlt-1, and PlGF were measured by quantitative sandwich ELISA using species-specific commercial kits from R&D System Inc. (Minneapolis, MN).

### Liquid chromatography-tandem mass spectrometric (LC/MS/MS) analysis

Serum concentrations of 16 Cers and 10 DHCers were measured by liquid chromatography-tandem mass spectrometric assay. The measurements were implemented for sera by the analytical methodology as previously described (55).

### Statistical analyses

The differentiating power of each gene in non-PE and PE placental tissues from multiple GEO datasets was combined by meta-analysis. The optimal Cer marker in women’s serum was determined by Mann–Whitney U test P-value, fold change, and AUC in the testing cohort. Performance of Lep, the optimal Cer marker, and Lep/Cer ratio in predicting PE was measured in the validation cohort by calculating AUC, sensitivity, specificity, PPV, and NPV. A time-to-event analysis was performed to calculate the gap between the time of prediction and the time of confirmatory diagnosis of PE. Results were compared with a reference point sFlt-1/PlGF ratio. Statistical analyses were preformed using R packages (56).

## RESULTS

### Meta-analysis confirmed the significant elevation of placental Lep expression in PE placentas

To test the ability of placental Lep levels to differentiate non-PE from PE pregnancies, we performed a meta-analysis of gene expression profiles based on six PE placental studies (GSE4707, GSE10588, GSE24129, GSE25906, GSE44711, and GSE54618; see Table I). Among the 13,137 genes, Lep in PE placentas had the maximal change (1.9-fold) and the most significant difference (*P* < 0.0001) compared with non-PE placentas (Fig. 2 and Appendix 1). Furthermore, in each study, Lep levels were higher in PE compared with non-PE placentas (*P* < 0.05; Fig. 3). The overexpression of the Lep transcripts was found to be significant in all PE women (including early-onset, late-onset, and severe PE).

**Table I.** Microarray datasets for meta-analysis on leptin (Lep) levels in placentas from non-preeclampsia (PE) and PE women.

| **Study** | **Non-PE, n** | **PE, n** |
| --- | --- | --- |
| **GSE4707** | 4 | 5 early-onset, 5 late-onset |
| **GSE10588** | 26 | 17 severe PE |
| **GSE24129** | 8 | 8 |
| **GSE25906** | 37 | 23 |
| **GSE44711** | 8 | 8 early-onset |
| **GSE54618** | 12 | 12 |
| **Total** | 95 | 78 |

**Figure 2.**
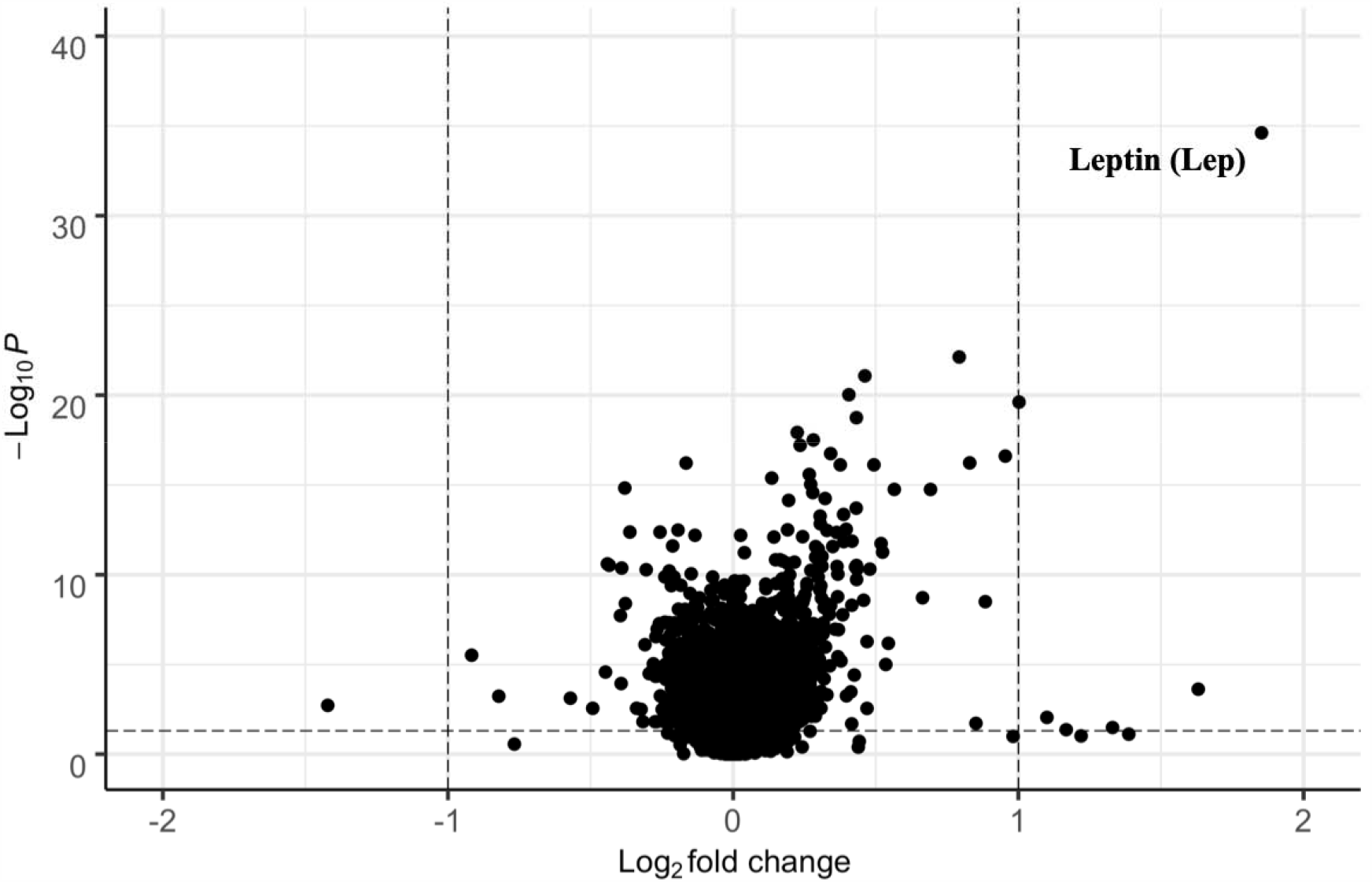
Meta-analysis identified differentially expressed genes in placentas from preeclamptic (PE) women. Volcano plot, fold changes (FC) on the X-axis and –log(*P*) on the Y-axis, was used to evaluate the performance of each placental gene that differentiates PE from non-preeclamptic (non-PE) women.

**Figure 3.**
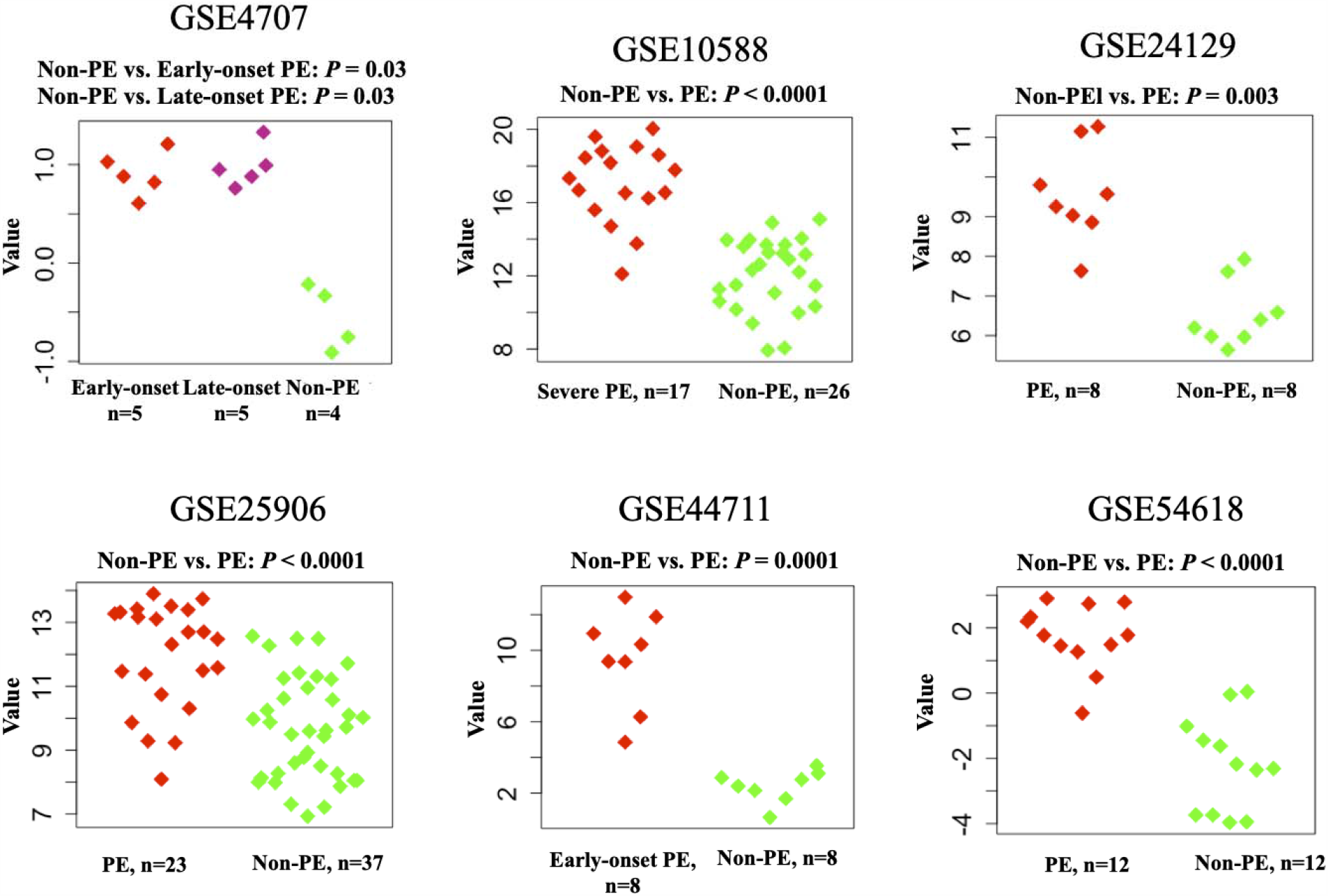
Transcriptional quantification of leptin (Lep) genes: a comparison between non-preeclamptic (PE) and preeclamptic (PE) placental expressions at delivery.

### PubMed meta-analysis identified Cer as the downstream metabolic messenger of Lep cascade

To discover the potential metabolic messengers downstream of Lep signaling cascade, we conducted a literature mining study based on the PubMed database. With the keyword of “Leptin; Lipid Metabolism”, 4212 publications were obtained from years of 2000 to 2020. Among 4212 publications, 78 studies were designed to investigate the biology underlying the cytoplasmic interactions between Lep and Cer at the molecular level. Afterwards, 14 and 150 publications were obtained by searching the keywords of “Ceramide; Pregnancy” and “Ceramide; Preeclampsia”, respectively, suggesting potential mechanistic implications of Cer in PE pathophysiology. Based on these results, Cer and its biosynthetic precursor dihydroceramide (DHCer) were selected as the biomarker candidates to generate our hypothesis and launched the following analyses.

### Serological quantitative analysis confirmed Lep, Cer, and DHCer as maternal markers for PE at diagnosis

We measured levels of circulating Lep in women from the testing cohort (testing cohort included 7 non-PE and 8 PE women; demographic data are shown in Table II). Concentrations of Lep were significantly higher in PE women than in women without PE between 25 and 31 wks’ of gestation (*P* = 0.02, 2.97-fold; Fig. 4A, Appendices 2 and 3).

**Table II.** Demographics of Cohort I. GA: gestational age.

| Characteristic | Non-PE | Preeclampsia (PE) |
| --- | --- | --- |
|  | n = 8 (53.3%) | n = 7 (46.7 %) |
| **Ethnicity** |  |  |
| African-American | 1 (12.5%) | 2 (28.6%) |
| Asian | 0 (0%) | 2 (28.6%) |
| Hispanic | 6 (75.0%) | 3 (42.8%) |
| Other | 1 (12.5%) | 0 (0%) |
| Age (years) | 27.0±4.9 | 23.1±5.8 |
| Wks' GA<sup>a</sup> | 28.9±2.0 | 27.8±1.8 |
| **a – Time of blood collection** |  |  |
| **GA – Gestational age** |  |  |

**Figure 4.**
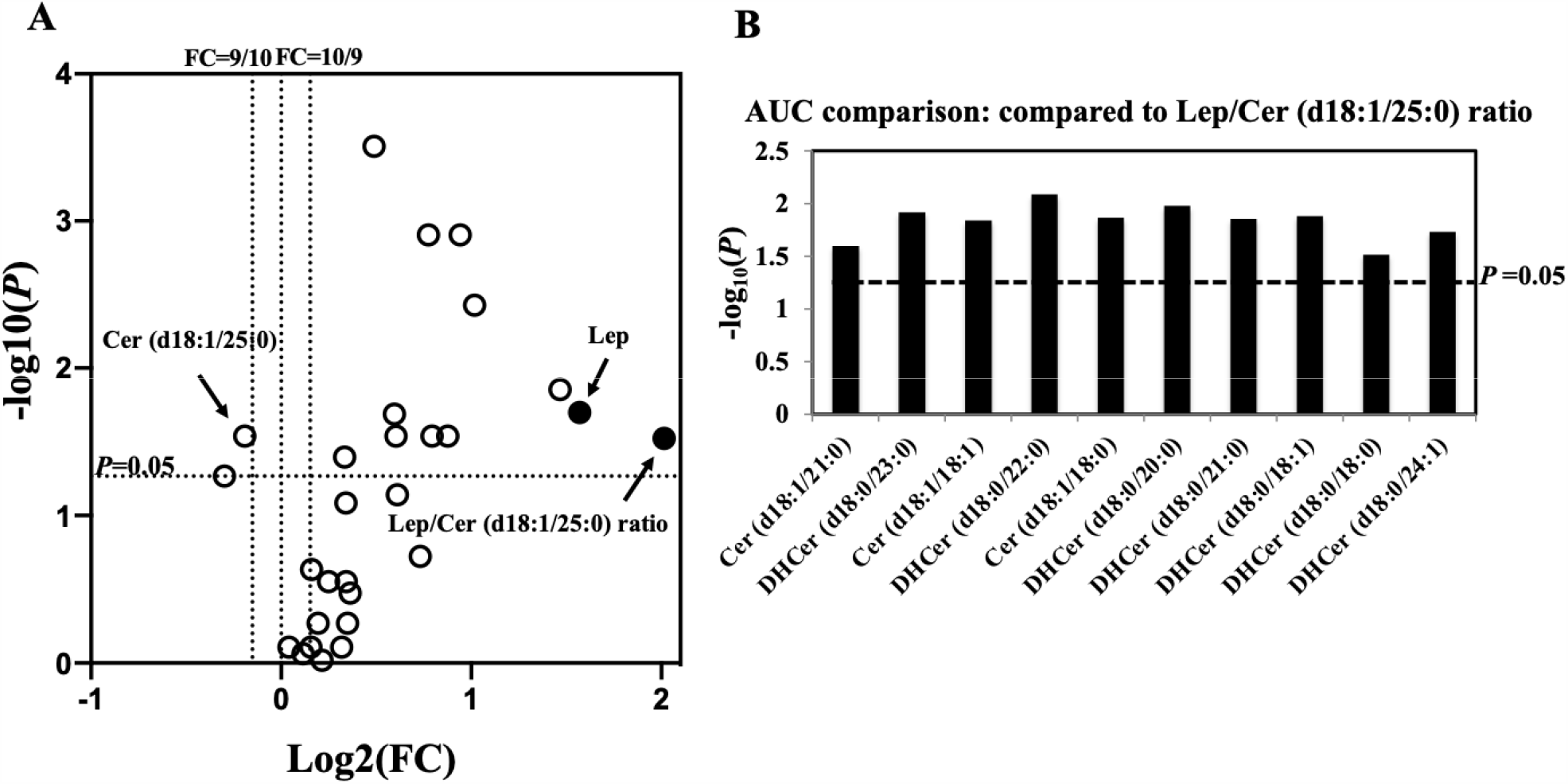
Comprehensive mass spectrometric analyses of 26 Cers/DHCers for preeclampsia (PE) diagnosis in the testing cohort. A: Fold change of each analyte between non-preeclamptic (non-PE) and preeclamptic (PE) women. A total of 11 Cers had *P* < 0.05. B: Area under curve (AUC) comparison between Lep/Cer(d18:1/25:0) ratio and other Lep-Cer combinations using each of the significant Cers in conjunction with Lep. DeLong test *P* were calculated (the y-axis). Lep: leptin. Cer: ceramide. DHCer: dihydroceramide.

We also characterized gestational profiles of 16 Cers and 10 DHCers in maternal sera (Fig. 4, Appendices 2 and 3). A total of 11 candidates (4 Cers and 7 DHCers) were significantly altered in PE (*P* < 0.05; Fig. 4A). Among the 11 candidates, Cer (d18:1/25:0) was found to be the only Cer that significantly changed compared with Lep in PE women (0.88-fold).

### Lep/Cer (d18:1/25:0) ratio differentiated PE from normal pregnancies

The Lep/Cer (d18:1/25:0) ratio had the largest area under the curve (AUC) compared with other marker combinations (Fig. 4B and Appendix 4) for differentiating PE from non-PE women in the testing cohort. Furthermore, compared with the individual Lep and Cer (d18:1/25:0) levels, the Lep/Cer (d18:1/25:0) ratio showed a lower *P* (0.006 vs 0.02 and 0.03, respectively), a larger fold change (4.04 vs 2.97 and 0.88, respectively), and a higher AUC (0.911 vs 0.875 and 0.839, respectively; Table III and Appendix 5).

**Table III.** Maternal serum leptin (Lep)/Cer (d18:1/25:0) ratio at 25–31 wks’ GA is a strong marker of preeclampsia (PE) in Cohort I.

| Marker | *P*<sup>a</sup> | Fold Change | AUC<sup>b</sup> |
| --- | --- | --- | --- |
| Lep | 0.02 | 2.97 | 0.875 |
| Cer<sup>c</sup> (d18:1/25:0) | 0.03 | 0.88 | 0.839 |
| Lep/Cer (d18:1/25:0) ratio | 0.006 | 4.04 | 0.911 |

<sup>a</sup>Mann–Whitney U test
<sup>b</sup>AUC: area under the curve
<sup>c</sup>Cer: ceramide

### Validation analysis found that Lep/Cer (d18:1/25:0) ratio predicts PE early in gestation

When we evaluated the predictive performance of the Lep/Cer (d18:1/25:0) ratio in the validation cohort (20 women without PE with 55 samples, and 20 women with PE with 51 samples; see Table IV and Fig. 5), We found that among the 20 PE women, 5 and 13 had early- and late-onset PE, respectively. The dates of diagnosis were missing in the remaining 2 women.

**Table IV.** Demographics of Cohort II. GA: gestational age. PE: Preeclampsia.

| **Characteristic** | **Non-PE (n = 20)** | **PE (n = 20)** |
| --- | --- | --- |
| **Race, n (%)** |  |  |
| **White** | 20 (100) | 9 (45) |
| **Asian** | 0 (0) | 5 (25) |
| **African American** | 0 (0) | 1 (5) |
| **Other** | 0 (0) | 5 (25) |
| **Age, years** | 31.9±4.8 | 31.8±6.0 |
| **GA at delivery, wks** | 39.5±1.2 | 36.7±3.3 |
| **Early-onset PE (Diagnosed < 34 wks' GA), n (%)** | NA | 5 (25) |
| **Diagnosed with severe PE, n (%)** | NA | 10 (50) |
| **GA – Gestational age** |  |  |

**Figure 5.**
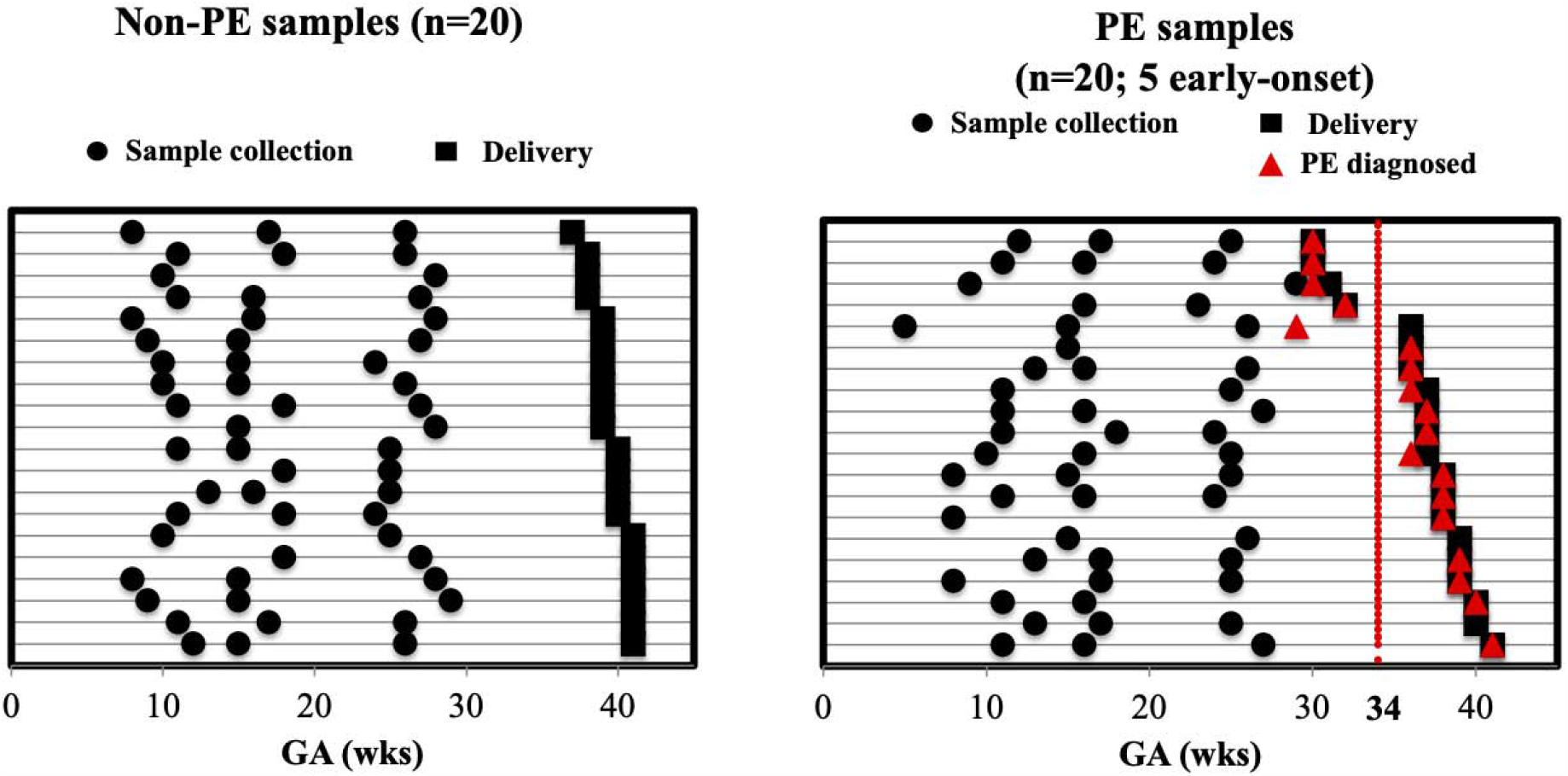
Sample collection timelines from the validation cohort: Serial blood sampling from each non-preeclamptic (PE) and preeclamptic (PE) woman at different gestational ages (GAs). Times of sample collection, delivery, and confirmatory PE diagnosis of each woman (denoted by each row) are represented by black circles, black squares, and red-filled triangles, respectively.

We observed an increase in serum Lep (*P* < 0.0001; 1.71-fold) and a decrease in Cer (d18:1/25:0) (*P* < 0.0001; 0.70-fold) in PE women at 5 to 29 wks’ of gestation (Appendix 6), which resulted in an elevated Lep/Cer (d18:1/25:0) ratio (Fig. 6). Most notably, this *ratio* was a significantly better predictor of all types of PE (AUC = 0.887) than Lep (AUC = 0.809; *P* = 0.0006) or Cer (d18:1/25:0) (AUC = 0.790; *P* = 0.008) levels alone. In addition, the ratio performed well at wider gestational windows from 5 to 15 and 16 to 29 wks (AUC = 0.876 and 0.892, respectively) than the individual markers: Lep (AUC = 0.868; *P* = 0.4 and 0.824; *P* = 0.1, respectively) and Cer (d18:1/25:0) (AUC =0.868; *P* = 0.1 and 0.747; *P* = 0.02, respectively). These results were consistent with the univariate and multivariate results found in non-obese women (Appendix 7). The Lep/Cer (d18:1/25:0) ratio accounted for 31% of the variation in the PE outcome, higher than that with Lep (15%), Cer (d18:1/25:0) (15%), and pre-pregnancy body mass index (BMI) (8%). In a multivariate analysis, the explained variance of Lep was dominant early in gestation (5 to 15 wks), while Cer (d18:1/25:0) was dominant in mid-gestation (16 to 29 wks; Appendix 7B). The longitudinal profiling of Lep and Cer (d18:1/25:0) levels improved the predictive performance of Lep/Cer (d18:1/25:0) ratio. Moreover, the Lep/Cer (d18:1/25:0) ratio outperformed the sFlt-1/PlGF ratio in predicting impending PE, with a lower *P* (Fig. 6) and a higher AUC (*P* < 0.0001; Appendix 6D).

**Figure 6.**
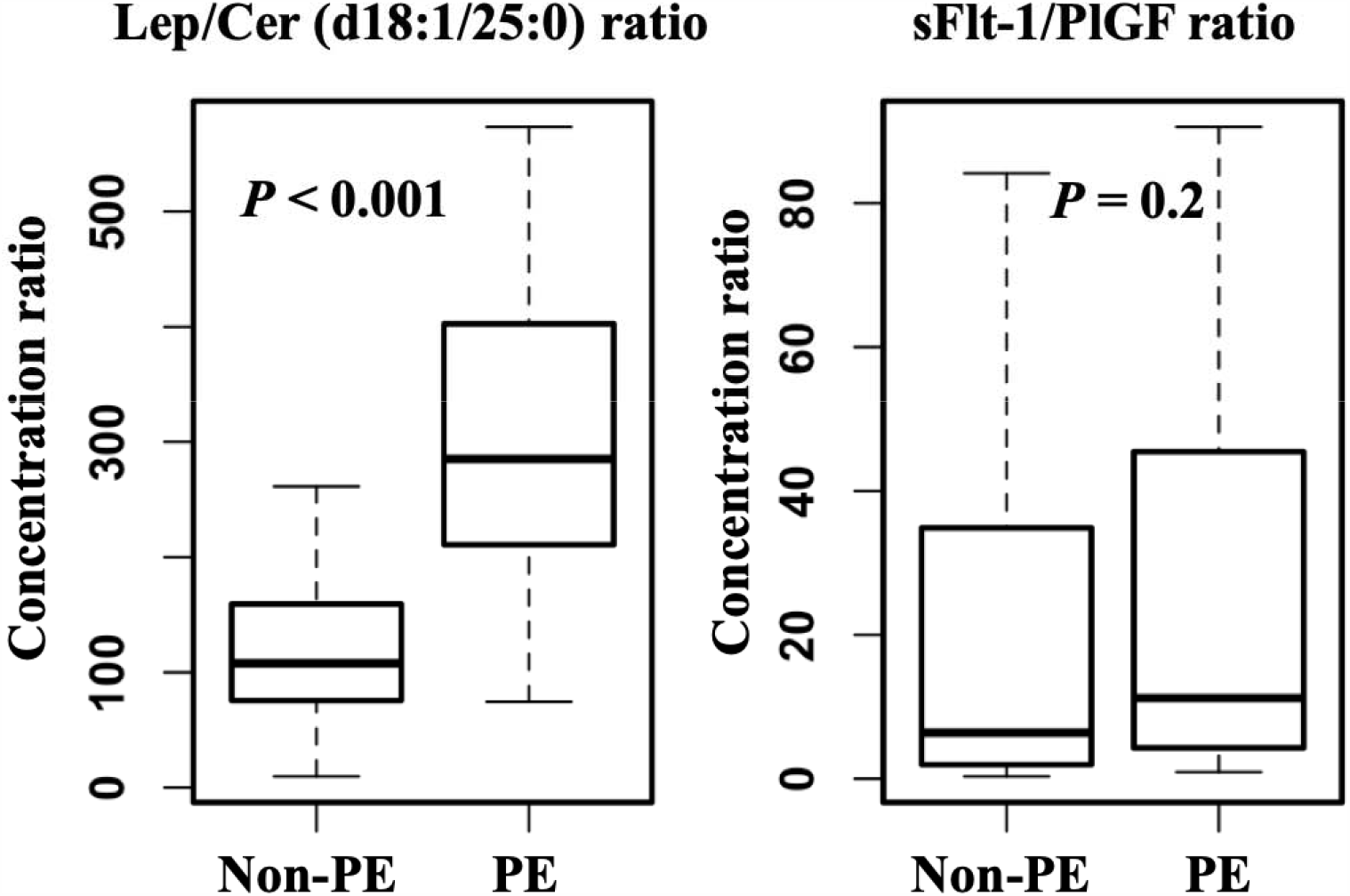
Comparisons of maternal serum levels between non-preeclamptic (PE) and preeclamptic (PE) pregnancies in the validation cohort. Left: Lep/Cer (d18:1/25:0) ratio; Right: sFLT-1/PlGF ratio. Lep: leptin. Cer: ceramide.

Time-to-event analysis at 5 to 25 wks (Fig. 7) compared the Lep/Cer (d18:1/25:0) ratio and the sFlt-1/PlGF ratio in predicting the impending PE. Among the 18 PE women with known diagnoses dates, 83% (15/18; 5 early-onset and 10 late-onset) were identified by the Lep/Cer (d18:1/25:0) ratio 11 or more weeks prior to their clinical diagnosis. In contrast, the sFlt-1/PlGF ratio only identified 22% (4/18; 1 early-onset and 3 late-onset) of PE women 11 or more weeks prior to the diagnosis. The Lep/Cer (d18:1/25:0) ratio was able to predict impending PE a median of 23.0 [95% confidence interval (CI): 12.8, 30.7] wks prior to the confirmatory diagnosis.

**Figure 7.**
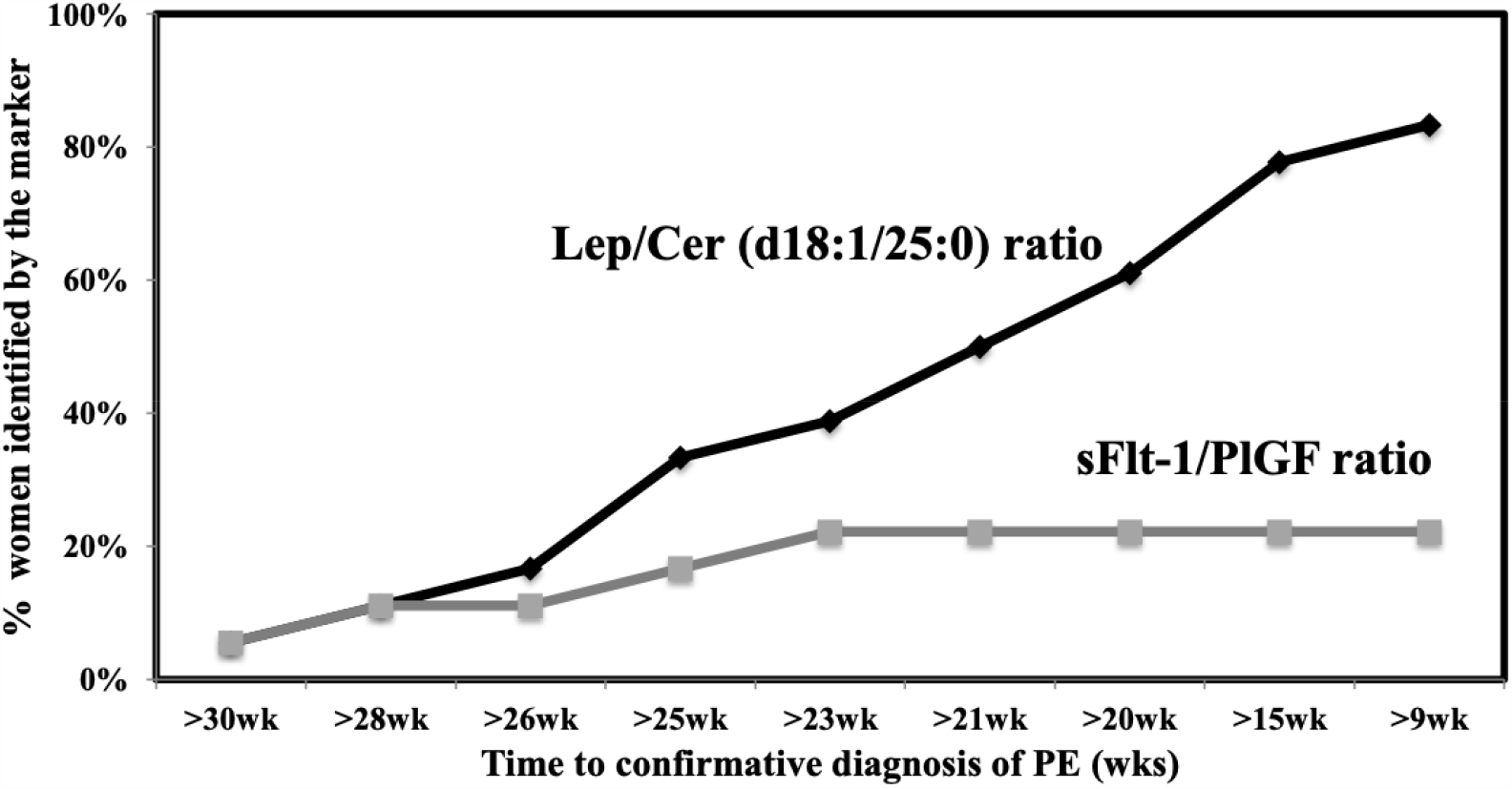
Comparative analysis between the ratios of Lep/Cer (d18:1/25:0) and sFLT-1/PlGF in predicting impending preeclampsia (PE). X-axis: the duration of time (wks) from the sampling to PE confirmatory diagnosis. Y-axis: the percentage of the PE women who were identified as high-risk within the specified duration before a confirmatory diagnosis. Lep: leptin. Cer: ceramide. wks: weeks.

As shown in Fig. 8A, the Lep/Cer (d18:1/25:0) ratio correctly classified 85% (17/20) of women with impending PE and 90% (18/20) of pregnancies without PE at 5 to 25 wks, giving a sensitivity of 85% (17/20), a specificity of 90% (18/20), a positive predictive value (PPV) of 89% (17/19), and a negative predictive value (NPV) of 86% (18/21). In contrast, 40% (8/20) of women with subsequent PE and 45% (9/20) of women without PE were correctly classified by the sFlt-1/PlGF ratio, yielding a sensitivity of only 40% (8/20), a specificity of 45% (9/20), a PPV of 42% (8/19), and a NPV of 43% (9/21) (Fig. 8B). In addition, the Lep/Cer (d18:1/25:0) ratio had a higher AUC than the sFlt-1/PlGF ratio at 5 to 25 wks [0.92 (95% CI: 0.86, 0.98) vs. 0.52 (95% CI: 0.39, 0.64); *P* < 0.001].

**Figure 8.**
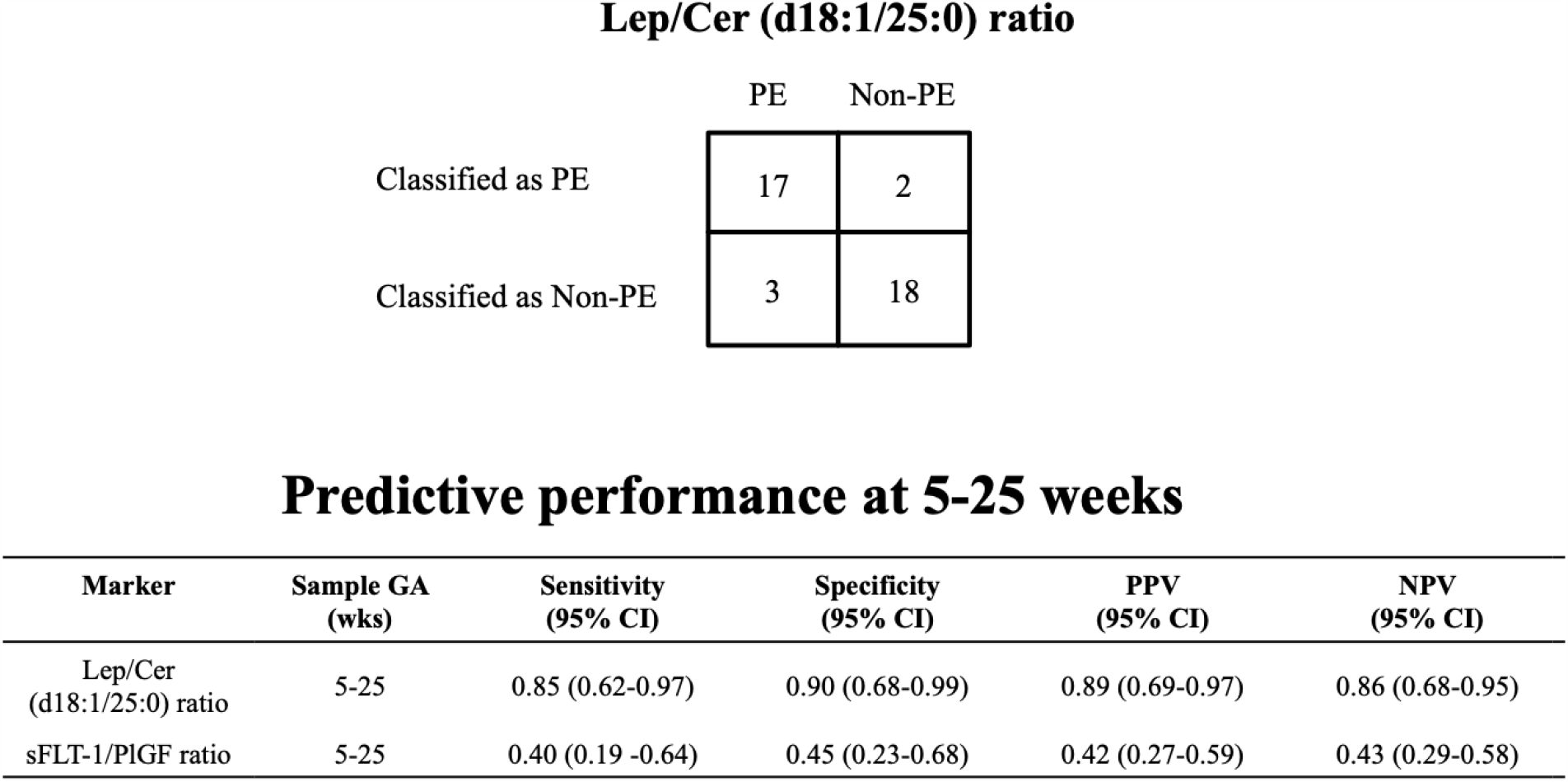
Individual-level performance of the Lep/Cer (d18:1/25:0) ratio in predicting impending preeclampsia (PE). A: 2×2 table. B: Sensitivity, specificity, positive predictive value (PPV), and negative predictive value (NPV). Lep: leptin. CI: confidence interval. GA: gestational age. Cer: ceramide.

## DISCUSSION

Early prediction of PE remains a challenge in current clinical practice. Known traditional risk factors inadequately identify women who will develop PE early in gestation (12–14,21,22). To aim the discovery of novel serological markers with better predictive power for PE at early gestations, we applied a multi-omics approach, integrating differentially expressed gene (DEGs) from placental mRNA expression multiplex analysis and lipidomic database mining of existing literature, to identify novel PE biomarkers, which are Lep and Cer. By characterizing the maternal serological profiles of Lep and Cer using commercial ELISA and reported liquid chromatography-tandem mass spectrometry (LC/MS/MS) assays, we validated the up-regulated Lep and down-regulated Cer(d18:1/25:0) in a case-control testing cohort with maternal sera collected at confirmative diagnosis of PE. With a cohort of longitudinally collected maternal sera from both PE and non-PE women, we further assessed the PE-predictive power of the Lep/Cer (18:1/25:0) ratio early in gestation. Our results validated this ratio as a better serological predictor of impending PE than the established sFlt-1/PlGF ratio. Our findings demonstrated that the use of the Lep/Cer (d18:1/25:0) ratio can identify women at high risk of developing PE at a substantially earlier time window during pregnancy (at 5 to 25 wks) than the sFlt-1/PlGF ratio (after 25 wks). Moreover, our results showed that, compared with the sFlt-1/PlGF ratio, the Lep/Cer (d18:1/25:0) ratio has better sensitivity (40% vs 85%), specificity (45% vs 90%), PPV (42% vs 89%), and NPV (43% vs 86%). Therefore, early in gestation, the Lep/Cer (d18:1/25:0) ratio outperforms the established sFlt-1/PlGF ratio and is a predictor of impending PE. The fact that the Lep/Cer (d18:1/25:0) ratio increases early in gestation in pregnant women who later develop PE offers an opportunity for predicting PE prior to the onset of clinical signs and symptoms. Integration of the ratio into a high-risk screening tool might allow patient identification at a pre-symptomatic stage. In addition, the concept of integrating a transcriptomic approach in placenta tissue with a lipidomic approach in serum is novel, as it combines the merits of studies in tissue whose focuses are more towards the pathogenesis and pathophysiology with those study in serum whose focuses are more towards the clinical translation. Taking the candidates obtained from the discovery phase to the validation phase makes the findings of this study translatable into clinical practice.

Previous studies have suggested that placental trophoblast cells are a leading source of circulating Lep in pregnancy (57), where Lep increases progressively in the first and second trimesters, peaks in the third, and returns to pre-pregnancy levels prior to parturition (58,59). In early gestation, Lep may play a critical role in modulating essential biological activities such as proliferation, protein synthesis, invasion, and apoptosis of trophoblast cells (60,61). Failure of trophoblastic invasion might result in incomplete remodeling of the maternal spiral arteries and inadequate placental perfusion to the embryo (62–64), leading to various disorders of reproduction and gestation such as intrauterine growth restriction (65), PE (66), gestational diabetes mellitus (67), and recurrent miscarriage (68). Other recent studies documented significant elevations of Lep expression in preeclamptic placentas (69–73). In the current study, we found a significant upregulation of Lep in maternal sera of PE women, which is consistent with previous reports (68–72).

The gestational dysregulation of Cer metabolism is believed to induce the aberrant de novo synthesis and lysosomal breakdown of Cer, which leads to trophoblast cell autophagy, dysfunctional development of placenta, and eventually pregnancy complications like PE (45,74,75). Cer (d18:1/25:0) is an unusual odd-chain species of the Cer family that is generated by de novo synthesis based on 25:0 fatty acid. Such odd-chain fatty acids are mainly from dairy products and meat from ruminant animals (76). Cer (d18:1/25:0) was previously described as a potential urinary marker of inflammation-induced alcoholic liver disease (77). Recently, an elevation of the Cer (d18:1/25:0) was also identified as a serum prognostic marker to predict various acute diseases, including cardiovascular death, myocardial infarction, and stroke in patients with acute myocardial infarction within an ensuing 12-month period (78). Our results suggested the pathological implications of Cer (d18:1/25:0) in the development of pregnancy complicated by PE, which might provide additional insights into the mechanistic roles of Cer (d18:1/25:0) and other odd-chain Cer species in PE pathophysiology.

An association between Lep and Cer has been reported in several studies. Lep was shown to exert its anorexigenic action by promoting mitochondrial lipid oxidation in both adipose and non-adipose tissues to alleviate ectopic accumulation of lipotoxic Cer via the activation of AMP-activated protein kinase (AMPKK/AMPK) (79,80). The de novo synthesis of Cer was found to play prominent roles in modulating downstream signaling of central Lep’s activity via mediation of malonyl-CoA, carnitine palmitoyl transferase-1c, and serine palmitoyl transferase (39–41). Persistent elevation of circulating Lep also appears to induce resistance at the level of the Lep receptor, which accounts for attenuated potency of Lep to alleviate the accumulated cytotoxic Cer (39–41). Our data suggest a crosstalk between Lep’s activity and de novo Cer synthesis. Lep functions well as a predictor of PE early in gestation, while Cer (d18:1/25:0) performs better at mid-gestation. The Lep/Cer (d18:1/25:0) ratio has a better predictive performance than Lep or Cer (d18:1/25:0) levels alone. Our findings revealed a correlation between the biological patterns of the two markers during PE progression, which might add value to existing knowledge about the Lep-Cer relationships.

Our study has several limitations. First, the sample sizes of the cohorts were small and lacked racial heterogeneity–thus, the generalizability of the results awaits larger and more racially diverse study populations. Second, longitudinal collections of blood samples were not evenly distributed over gestation. Finally, we did not investigate the exact tissue of origin, where Lep is overexpressed in PE women. By using a conditional knock-in placental Lep transgenic mouse model, it may be possible to elucidate the mechanistic role of placental Lep in the pathogenesis of PE early in pregnancy.

## CONCLUSIONS

The disruptions of gestational homeostasis involving placenta-related biological networks are important factors contributing to the pathophysiology of PE. Lep, an endocrine regulator of body energy repletion, and Cer (d18:1/25:0), a bioactive metabolic messenger downstream of Lep, were identified by the multi-omics discovery to be significantly up- and down-regulated in the maternal circulation of women with PE. The Lep/Cer (d18:1/25:0) ratio was demonstrated to provide augmented predictive power in differentiating PE from a pregnancy without PE before a confirmatory diagnosis can be made. The maternal Lep/Cer (d18:1/25:0) ratio, with an earlier elevation in gestation, is a superior prognostic marker than the sFlt-1/PlGF ratio. If validated as a laboratory developed test or in vitro diagnostics, the deployment of the Lep/Cer ratio test to assess PE and proactively manage asymptomatic early pregnancies should have profound impact on PE care.

## Data Availability

The datasets used and/or analyzed in this study are available upon request to the corresponding author.

## List of Abbreviation

ACC: Acetyl-CoA carboxylase
AUC: Area Under the Curve
AMPKK/AMPK: AMP-activated protein kinase
BMI: Body Mass Index
Cer: Ceramide
CI: Confidence Interval
DHCer: Dihydroceramide
ELISA: Enzyme-Linked Immunosorbent Assay
GEO: Gene Expression Omnibus
Lep: Leptin
LC/MS/MS: Liquid Chromatography-Tandem Mass Spectrometry
NPV: Negative Predictive Value
PlGF: Placental Growth Factor
PPV: Positive Predictive Value
PE: Preeclampsia
sFlt-1: Soluble fms-Like Tyrosine Kinase
Wk: Week

## Acknowledgements

We thank our colleagues in the March of the Dimes Prematurity Research Center at Stanford University and Pediatrics Proteomics Group for critical discussions.

## Author contributions

XBL, KGS, and HJC contributed to concept development and design. JY, RJW, and DKS contributed to the acquisition of data.

QH, SH, XY, JY, ZL, JS, ZL, ST, WL, XZ, LM, SL, RJW, GMS, DKS, JCW, and DBM contributed to the analysis and interpretation of data.

QH and SH drafted the manuscript.

XY, JY, ZL, JS, ZL, ST, WL, XZ, LM, SL, RJW, GMS, DKS, HJC, JCW, DBM, KGS, and XBL critically revised the manuscript.

All the authors gave final approval of the version to be submitted and agreed to be accountable for all aspects of the work.

## Reference

1. Sibai B, Dekker G, Kupferminc M. Pre-eclampsia. Lancet (London, England). 2005 Feb;365(9461):785–99.

2. Noris M, Perico N, Remuzzi G. Mechanisms of disease: Pre-eclampsia. Nature clinical practice Nephrology. 2005 Dec;1(2):98–114; quiz 120.

3. Roberts JM, Pearson GD, Cutler JA, Lindheimer MD. Summary of the NHLBI Working Group on Research on Hypertension During Pregnancy. Hypertension in pregnancy. 2003;22(2):109–27.

4. Roberts JM, Escudero C. The placenta in preeclampsia. Pregnancy hypertension. 2012 Apr;2(2):72–83.

5. Chaiworapongsa T, Chaemsaithong P, Yeo L, Romero R. Pre-eclampsia part 1: current understanding of its pathophysiology. Nature reviews Nephrology. 2014 Aug;10(8):466–80.

6. Ahmed A, Ramma W. Unravelling the theories of pre-eclampsia: are the protective pathways the new paradigm? British journal of pharmacology. 2015 Mar;172(6):1574–86.

7. ACOG committee opinion. Antenatal corticosteroid therapy for fetal maturation. American College of Obstetricians and Gynecologists. International journal of gynaecology and obstetrics: the official organ of the International Federation of Gynaecology and Obstetrics. 2002 Jul;78(1):95–7.

8. Vatten LJ, Nilsen TIL, Juul A, Jeansson S, Jenum PA, Eskild A. Changes in circulating level of IGF-I and IGF-binding protein-1 from the first to second trimester as predictors of preeclampsia. European journal of endocrinology. 2008 Jan;158(1):101–5.

9. Raia-Barjat T, Prieux C, Gris J-C, Chapelle C, Laporte S, Chauleur C. Angiogenic factors for prediction of preeclampsia and intrauterine growth restriction onset in high-risk women: AngioPred study. The journal of maternal-fetal & neonatal medicine: the official journal of the European Association of Perinatal Medicine, the Federation of Asia and Oceania Perinatal Societies, the International Society of Perinatal Obstetricians. 2019 Jan;32(2):248–57.

10. Phipps EA, Thadhani R, Benzing T, Karumanchi SA. Pre-eclampsia: pathogenesis, novel diagnostics and therapies. Nature reviews Nephrology. 2019 May;15(5):275–89.

11. Armaly Z, Jadaon JE, Jabbour A, Abassi ZA. Preeclampsia: Novel Mechanisms and Potential Therapeutic Approaches. Frontiers in physiology. 2018;9:973.

12. Poon LC, Shennan A, Hyett JA, Kapur A, Hadar E, Divakar H, et al. The International Federation of Gynecology and Obstetrics (FIGO) initiative on pre-eclampsia: A pragmatic guide for first-trimester screening and prevention. International journal of gynaecology and obstetrics: the official organ of the International Federation of Gynaecology and Obstetrics. 2019 May;145 Suppl(Suppl 1):1–33.

13. Musa J, Mohammed C, Ocheke A, Kahansim M, Pam V, Daru P. Incidence and risk factors for pre-eclampsia in Jos Nigeria. African health sciences. 2018 Sep;18(3):584–95.

14. Chaiworapongsa T, Chaemsaithong P, Korzeniewski SJ, Yeo L, Romero R. Pre-eclampsia part 2: prediction, prevention and management. Nature reviews Nephrology. 2014 Sep;10(9):531–40.

15. Rodriguez-Lopez M, Wagner P, Perez-Vicente R, Crispi F, Merlo J. Revisiting the discriminatory accuracy of traditional risk factors in preeclampsia screening. Spracklen CN, editor. PLOS ONE [Internet]. 2017 May 25 [cited 2020 Jun 18];12(5):e0178528. Available from: https://dx.plos.org/10.1371/journal.pone.0178528

16. Kopcow HD, Karumanchi SA. Angiogenic factors and natural killer (NK) cells in the pathogenesis of preeclampsia. Journal of reproductive immunology. 2007 Dec;76(1–2):23–9.

17. Zhou CC, Ahmad S, Mi T, Xia L, Abbasi S, Hewett PW, et al. Angiotensin II induces soluble fms-Like tyrosine kinase-1 release via calcineurin signaling pathway in pregnancy. Circulation research. 2007 Jan;100(1):88–95.

18. Kim S-Y, Ryu H-M, Yang J-H, Kim M-Y, Han J-Y, Kim J-O, et al. Increased sFlt-1 to PlGF ratio in women who subsequently develop preeclampsia. Journal of Korean medical science. 2007 Oct;22(5):873–7.

19. Romero R, Nien JK, Espinoza J, Todem D, Fu W, Chung H, et al. A longitudinal study of angiogenic (placental growth factor) and anti-angiogenic (soluble endoglin and soluble vascular endothelial growth factor receptor-1) factors in normal pregnancy and patients destined to develop preeclampsia and deliver a small fo. The journal of maternal-fetal & neonatal medicine: the official journal of the European Association of Perinatal Medicine, the Federation of Asia and Oceania Perinatal Societies, the International Society of Perinatal Obstetricians. 2008 Jan;21(1):9–23.

20. Carty DM, Delles C, Dominiczak AF. Novel biomarkers for predicting preeclampsia. Trends in cardiovascular medicine. 2008 Jul;18(5):186–94.

21. Verlohren S, Galindo A, Schlembach D, Zeisler H, Herraiz I, Moertl MG, et al. An automated method for the determination of the sFlt-1/PIGF ratio in the assessment of preeclampsia. American journal of obstetrics and gynecology. 2010 Feb;202(2):161.e1-161.e11.

22. Hao S, You J, Chen L, Zhao H, Huang Y, Zheng L, et al. Changes in pregnancy-related serum biomarkers early in gestation are associated with later development of preeclampsia. PloS one. 2020;15(3):e0230000.

23. Kelly RS, Croteau-Chonka DC, Dahlin A, Mirzakhani H, Wu AC, Wan ES, et al. Integration of metabolomic and transcriptomic networks in pregnant women reveals biological pathways and predictive signatures associated with preeclampsia. Metabolomics: Official journal of the Metabolomic Society. 2017 Jan;13(1).

24. Liu LY, Yang T, Ji J, Wen Q, Morgan AA, Jin B, et al. Integrating multiple “omics” analyses identifies serological protein biomarkers for preeclampsia. BMC medicine. 2013 Nov;11:236.

25. Aghaeepour N, Lehallier B, Baca Q, Ganio EA, Wong RJ, Ghaemi MS, et al. A proteomic clock of human pregnancy. American journal of obstetrics and gynecology. 2018 Mar;218(3):347.e1-347.e14.

26. Benny PA, Alakwaa FM, Schlueter RJ, Lassiter CB, Garmire LX. A review of omics approaches to study preeclampsia. Placenta [Internet]. 2020;92:17–27. Available from: http://www.sciencedirect.com/science/article/pii/S0143400420300187

27. Tarca AL, Romero R, Benshalom-Tirosh N, Than NG, Gudicha DW, Done B, et al. The prediction of early preeclampsia: Results from a longitudinal proteomics study. PloS one. 2019;14(6):e0217273.

28. Nguyen TPH, Patrick CJ, Parry LJ, Familari M. Using proteomics to advance the search for potential biomarkers for preeclampsia: A systematic review and meta-analysis. PloS one. 2019;14(4):e0214671.

29. Tarca AL, Romero R, Erez O, Gudicha DW, Than NG, Benshalom-Tirosh N, et al. Maternal whole blood mRNA signatures identify women at risk of early preeclampsia: a longitudinal study. The journal of maternal-fetal & neonatal medicine: the official journal of the European Association of Perinatal Medicine, the Federation of Asia and Oceania Perinatal Societies, the International Society of Perinatal Obstetricians. 2020 Jan;1–12.

30. Trifonova EA, Gabidulina T v, Ershov NI, Serebrova VN, Vorozhishcheva AY, Stepanov VA. Analysis of the placental tissue transcriptome of normal and preeclampsia complicated pregnancies. Acta naturae. 2014 Apr;6(2):71–83.

31. Harati-Sadegh M, Kohan L, Teimoori B, Mehrabani M, Salimi S. Analysis of polymorphisms, promoter methylation, and mRNA expression profile of maternal and placental P53 and P21 genes in preeclamptic and normotensive pregnant women. Journal of biomedical science. 2019 Nov;26(1):92.

32. Jung YW, Shim JI, Shim SH, Shin Y-J, Shim SH, Chang SW, et al. Global gene expression analysis of cellfree RNA in amniotic fluid from women destined to develop preeclampsia. Medicine. 2019 Jan;98(3):e13971.

33. Wu K, Liu F, Wu W, Chen Y, Zhang W. Bioinformatics approach reveals the critical role of TGF-beta signaling pathway in pre-eclampsia development. European journal of obstetrics, gynecology, and reproductive biology. 2019 Sep;240:130–8.

34. Friedman J. The long road to leptin. The Journal of clinical investigation. 2016 Dec;126(12):4727–34.

35. Masuzaki H, Ogawa Y, Sagawa N, Hosoda K, Matsumoto T, Mise H, et al. Nonadipose tissue production of leptin: Leptin as a novel placenta-derived hormone in humans. Nature Medicine [Internet]. 1997;3(9):1029–33. Available from: https://doi.org/10.1038/nm0997-1029

36. Pérez-Pérez A, Toro A, Vilariño-García T, Maymó J, Guadix P, Dueñas JL, et al. Leptin action in normal and pathological pregnancies. Vol. 22, Journal of Cellular and Molecular Medicine. Blackwell Publishing Inc.; 2018. p. 716–27.

37. Zhao H, Wong RJ, Kalish FS, Nayak NR, Stevenson DK. Effect of heme oxygenase-1 deficiency on placental development. Placenta. 2009 Oct;30(10):861–8.

38. Zhao H, Ozen M, Wong RJ, Stevenson DK. Heme oxygenase-1 in pregnancy and cancer: similarities in cellular invasion, cytoprotection, angiogenesis, and immunomodulation. Frontiers in pharmacology. 2014;5:295.

39. Gao S, Zhu G, Gao X, Wu D, Carrasco P, Casals N, et al. Important roles of brain-specific carnitine palmitoyltransferase and ceramide metabolism in leptin hypothalamic control of feeding. Proceedings of the National Academy of Sciences of the United States of America. 2011 Jun;108(23):9691–6.

40. Bonzon-Kulichenko E, Schwudke D, Gallardo N, Molto E, Fernandez-Agullo T, Shevchenko A, et al. Central leptin regulates total ceramide content and sterol regulatory element binding protein-1C proteolytic maturation in rat white adipose tissue. Endocrinology. 2009 Jan;150(1):169–78.

41. Unger RH, Roth MG. A new biology of diabetes revealed by leptin. Cell metabolism. 2015 Jan;21(1):15–20.

42. Huang Q, Hao S, Yao X, You J, Li X, Lai D, et al. High-throughput quantitation of serological ceramides/dihydroceramides by LC/MS/MS: pregnancy baseline biomarkers and potential metabolic messengers. Journal of Pharmaceutical and Biomedical Analysis [Internet]. 2020 Sep 22 [cited 2020 Sep 22];113639. Available from:]https://linkinghub.elsevier.com/retrieve/pii/S0731708520315259

43. Charkiewicz K, Goscik J, Blachnio-Zabielska A, Raba G, Sakowicz A, Kalinka J, et al. Sphingolipids as a new factor in the pathomechanism of preeclampsia - Mass spectrometry analysis. PLoS ONE. 2017 May 1;12(5).

44. del Gaudio I, Sasset L, Lorenzo A di, Wadsack C. Sphingolipid Signature of Human Feto-Placental Vasculature in Preeclampsia. International journal of molecular sciences. 2020 Feb;21(3).

45. Melland-Smith M, Ermini L, Chauvin S, Craig-Barnes H, Tagliaferro A, Todros T, et al. Disruption of sphingolipid metabolism augments ceramide-induced autophagy in preeclampsia. Autophagy. 2015;11(4):653– 69.

46. Nishizawa H, Pryor-Koishi K, Kato T, Kowa H, Kurahashi H, Udagawa Y. Microarray Analysis of Differentially Expressed Fetal Genes in Placental Tissue Derived from Early and Late Onset Severe Preeclampsia. Placenta. 2007 May 1;28(5–6):487–97.

47. Sitras V, Paulssen RH, Grønaas H, Leirvik J, Hanssen TA, Vårtun Å, et al. Differential Placental Gene Expression in Severe Preeclampsia. Placenta. 2009 May 1;30(5):424–33.

48. Nishizawa H, Ota S, Suzuki M, Kato T, Sekiya T, Kurahashi H, et al. Comparative gene expression profiling of placentas from patients with severe pre-eclampsia and unexplained fetal growth restriction [Internet]. Vol. 9, Reproductive Biology and Endocrinology. 2011 [cited 2020 Sep 30]. Available from: http://www.rbej.com/content/9/1/107

49. Tsai S, Hardison NE, James AH, Motsinger-Reif AA, Bischoff SR, Thames BH, et al. Transcriptional Profiling of Human Placentas from Pregnancies Complicated by Preeclampsia Reveals Disregulation of Sialic Acid Acetylesterase and Immune Signalling Pathways. Placenta. 2011;32(2):175–82.

50. Jebbink JM, Boot RG, Keijser R, Moerland PD, Aten J, Veenboer GJM, et al. Increased glucocerebrosidase expression and activity in preeclamptic placenta. Placenta. 2015 Feb 1;36(2):160–9.

51. Blair JD, Yuen RKC, Lim BK, McFadden DE, von Dadelszen P, Robinson WP. Widespread DNA hypomethylation at gene enhancer regions in placentas associated with early-onset pre-eclampsia. Molecular Human Reproduction [Internet]. 2013 Oct [cited 2020 Sep 30];19(10):697–708. Available from: /pmc/articles/PMC3779005/?report=abstract

52. Chen R, Sigdel TK, Li L, Kambham N, Dudley JT, Hsieh S, et al. Differentially Expressed RNA from Public Microarray Data Identifies Serum Protein Biomarkers for Cross-Organ Transplant Rejection and Other Conditions. Weiss ST, editor. PLoS Computational Biology [Internet]. 2010 Sep 23 [cited 2020 Sep 30];6(9):e1000940. Available from: https://dx.plos.org/10.1371/journal.pcbi.1000940

53. Morgan AA, Khatri P, Jones RH, Sarwal MM, Butte AJ. Comparison of multiplex meta analysis techniques for understanding the acute rejection of solid organ transplants From 2010 AMIA Summit on Translational Bioinformatics [Internet]. Vol. 11, BMC Bioinformatics. 2010 [cited 2020 Sep 30]. Available from: http://www.biomedcentral.com/1471-2105/11/S9/S6

54. Hypertension in Pregnancy [Internet]. Available from: https://journals.lww.com/greenjournal

55. Huang Q, Hao S, Yao X, You J, Li X, Lai D, et al. Quantitative LCMS for ceramides/dihydroceramides: pregnancy baseline biomarkers and potential metabolic messengers. bioRxiv [Internet]. 2020 Jan 1;2020.02.24.963462. Available from: http://biorxiv.org/content/early/2020/02/25/2020.02.24.963462.abstract

56. R Core Team. R: A language and environment for statistical computing. R Foundation for Statistical Computing, Vienna, Austria. URL: http://www.R-project.org/. 2014.

57. Masuzaki H, Ogawa Y, Sagawa N, Hosoda K, Matsumoto T, Mise H, et al. Nonadipose tissue production of leptin: leptin as a novel placenta-derived hormone in humans. Nature medicine. 1997 Sep;3(9):1029–33.

58. Bajoria R, Sooranna SR, Ward BS, Chatterjee R. Prospective function of placental leptin at maternal-fetal interface. Placenta. 2002;23(2–3):103–15.

59. Henson MC, Castracane VD. Leptin in pregnancy: an update. Biology of reproduction. 2006 Feb;74(2):218– 29.

60. Chehab FF. 20 years of leptin: leptin and reproduction: past milestones, present undertakings, and future endeavors. The Journal of endocrinology. 2014 Oct;223(1):T37–48.

61. Reitman ML, Bi S, Marcus-Samuels B, Gavrilova O. Leptin and its role in pregnancy and fetal development--an overview. Biochemical Society transactions. 2001 May;29(Pt 2):68–72.

62. Pollheimer J, Knofler M. Signalling pathways regulating the invasive differentiation of human trophoblasts: a review. Placenta. 2005 Apr;26 Suppl A:S21–30.

63. E Davies J, Pollheimer J, Yong HEJ, Kokkinos MI, Kalionis B, Knofler M, et al. Epithelial-mesenchymal transition during extravillous trophoblast differentiation. Cell adhesion & migration. 2016 May;10(3):310–21.

64. Staun-Ram E, Shalev E. Human trophoblast function during the implantation process. Reproductive biology and endocrinology: RB&E. 2005 Oct; 3:56.

65. Weiss G, Sundl M, Glasner A, Huppertz B, Moser G. The trophoblast plug during early pregnancy: a deeper insight. Histochemistry and cell biology. 2016 Dec;146(6):749–56.

66. Sheikh AM, Small HY, Currie G, Delles C. Systematic Review of Micro-RNA Expression in Pre-Eclampsia Identifies a Number of Common Pathways Associated with the Disease. PloS one. 2016;11(8): e0160808.

67. Perez-Perez A, Guadix P, Maymo J, Duenas JL, Varone C, Fernandez-Sanchez M, et al. Insulin and Leptin Signaling in Placenta from Gestational Diabetic Subjects. Hormone and metabolic research = Hormon- und Stoffwechselforschung = Hormones et metabolisme. 2016 Jan;48(1):62–9.

68. Serazin V, Duval F, Wainer R, Ravel C, Vialard F, Molina-Gomes D, et al. Are leptin and adiponectin involved in recurrent pregnancy loss? The journal of obstetrics and gynaecology research. 2018 Jun;44(6):1015–22.

69. Mise H, Sagawa N, Matsumoto T, Yura S, Nanno H, Itoh H, et al. Augmented placental production of leptin in preeclampsia: possible involvement of placental hypoxia. The Journal of clinical endocrinology and metabolism. 1998 Sep;83(9):3225–9.

70. el Shahat AM, Ahmed AB, Ahmed MR, Mohamed HS. Maternal serum leptin as a marker of preeclampsia. Archives of gynecology and obstetrics. 2013 Dec;288(6):1317–22.

71. Lacroix M, Battista M-C, Doyon M, Moreau J, Patenaude J, Guillemette L, et al. Higher maternal leptin levels at second trimester are associated with subsequent greater gestational weight gain in late pregnancy. BMC pregnancy and childbirth. 2016 Mar; 16: 62.

72. Taylor BD, Ness RB, Olsen J, Hougaard DM, Skogstrand K, Roberts JM, et al. Serum leptin measured in early pregnancy is higher in women with preeclampsia compared with normotensive pregnant women. Hypertension (Dallas, Tex: 1979). 2015 Mar;65(3):594–9.

73. Yeboah FA, Ngala RA, Bawah AT, Asare-Anane H, Alidu H, Hamid A-WM, et al. Adiposity and hyperleptinemia during the first trimester among pregnant women with preeclampsia. International journal of women’s health. 2017; 9: 449–54.

74. Bailey LJ, Alahari S, Tagliaferro A, Post M, Caniggia I. Augmented trophoblast cell death in preeclampsia can proceed via ceramide-mediated necroptosis. Cell death & disease. 2017 Feb;8(2): e2590.

75. Charkiewicz K, Goscik J, Blachnio-Zabielska A, Raba G, Sakowicz A, Kalinka J, et al. Sphingolipids as a new factor in the pathomechanism of preeclampsia - Mass spectrometry analysis. PloS one. 2017;12(5): e0177601.

76. Jenkins B, West JA, Koulman A. A review of odd-chain fatty acid metabolism and the role of pentadecanoic acid (C15:0) and heptadecanoic acid (C17:0) in health and disease. Vol. 20, Molecules. MDPI AG; 2015. p. 2425–44.

77. Sun H, Lv H, Zhang A, Wang X. Chapter 19 - Metabolic Biomarkers of Alcohol Liver Damage and the Intervention Effect of Yinchenhao Tang. In: Wang X, Zhang A, Sun HBT-C, editors. Boston: Academic Press; 2015. p. 293–304. Available from: http://www.sciencedirect.com/science/article/pii/B9780128031179000196

78. de Carvalho LP, Tan SH, Ow GS, Tang Z, Ching J, Kovalik JP, et al. Plasma Ceramides as Prognostic Biomarkers and Their Arterial and Myocardial Tissue Correlates in Acute Myocardial Infarction. JACC: Basic to Translational Science. 2018 Apr 1;3(2):163–75.

79. Minokoshi Y, Toda C, Okamoto S. Regulatory role of leptin in glucose and lipid metabolism in skeletal muscle. Indian journal of endocrinology and metabolism [Internet]. 2012 Dec;16(Suppl 3): S562–8. Available from: https://pubmed.ncbi.nlm.nih.gov/23565491

80. Turpin SM, Nicholls HT, Willmes DM, Mourier A, Brodesser S, Wunderlich CM, et al. Obesity-induced CerS6-dependent C16:0 ceramide production promotes weight gain and glucose intolerance. Cell Metabolism. 2014 Oct 7;20(4):678–86.

