## Supplemental materials for "Case finding of early pregnancies at risk of preeclampsia using maternal blood leptin/ceramide ratio: multi-omics discovery and validation from a longitudinal study"

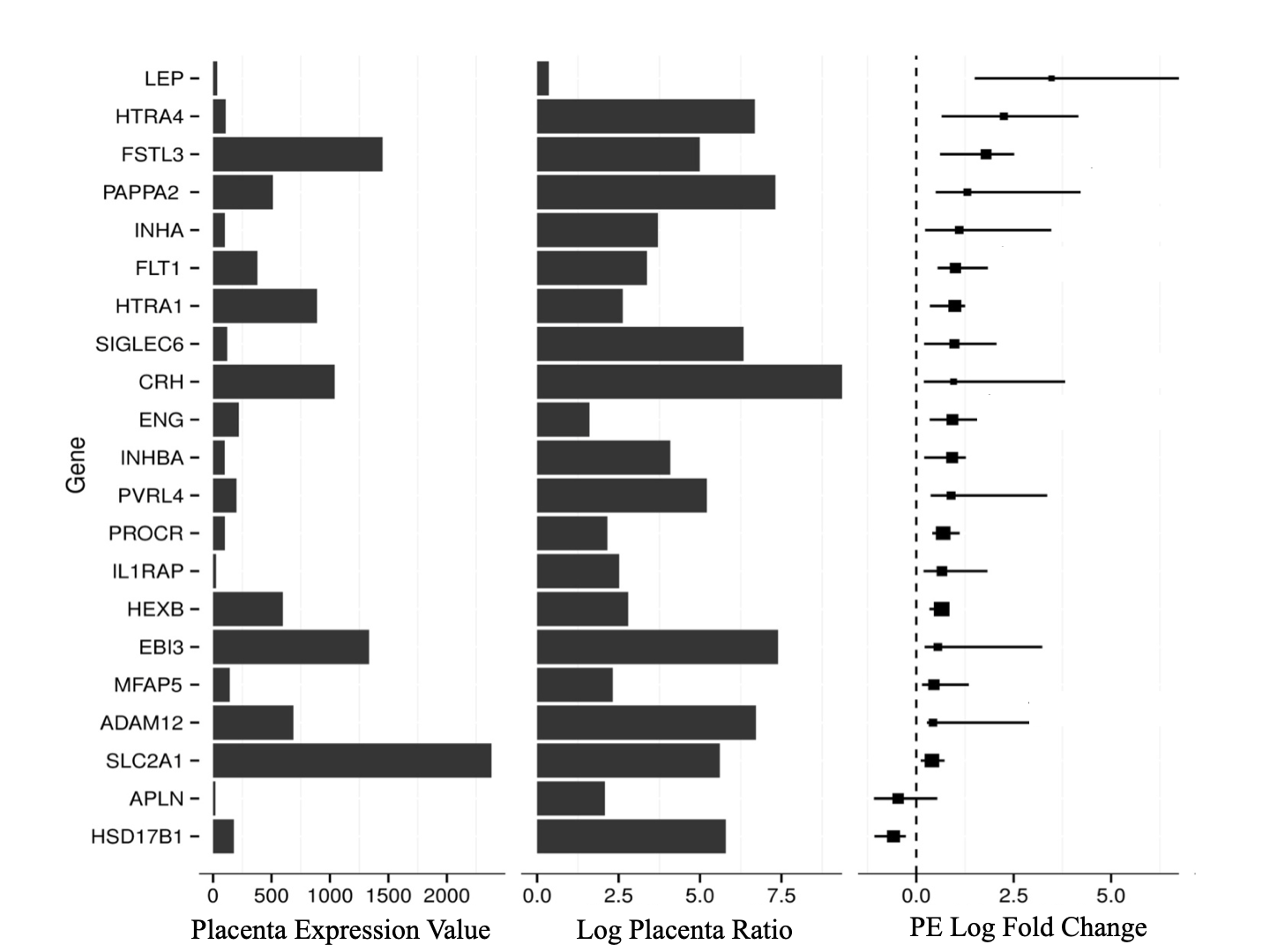

**Appendix 1.** Results of meta-analysis. Placenta expression value and placenta ratio: the expression values from the whole placental tissue to the average values from 15 other human tissues were compared and the significantly enriched genes are listed; PE log fold of change: global transcriptional profiling comparing non-preeclamptic (PE) to preeclamptic (PE) placentas. Leptin (Lep) is found to be the most differentiated transcript.

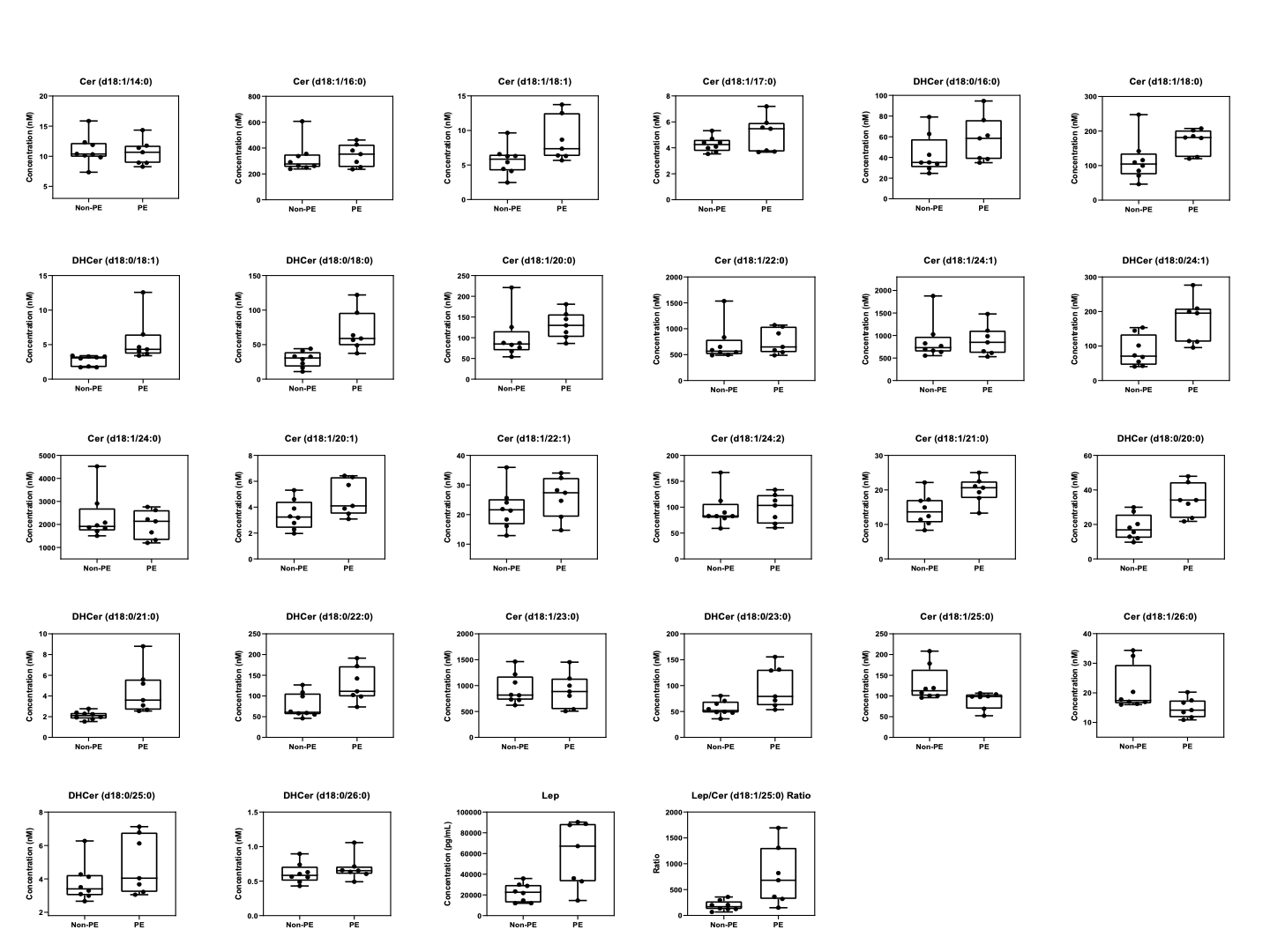

**Appendix 2.** These boxplots show maternal serum concentrations of each analyte in preeclamptic (PE) and non-preeclamptic (non-PE) women in the testing cohort. Analytes include 24 Cers/DHCers, Lep, and the ratio of Lep/Cer (d18:1/25:0). Lep: leptin. Cer: ceramide. DHCer: dihydroceramide.

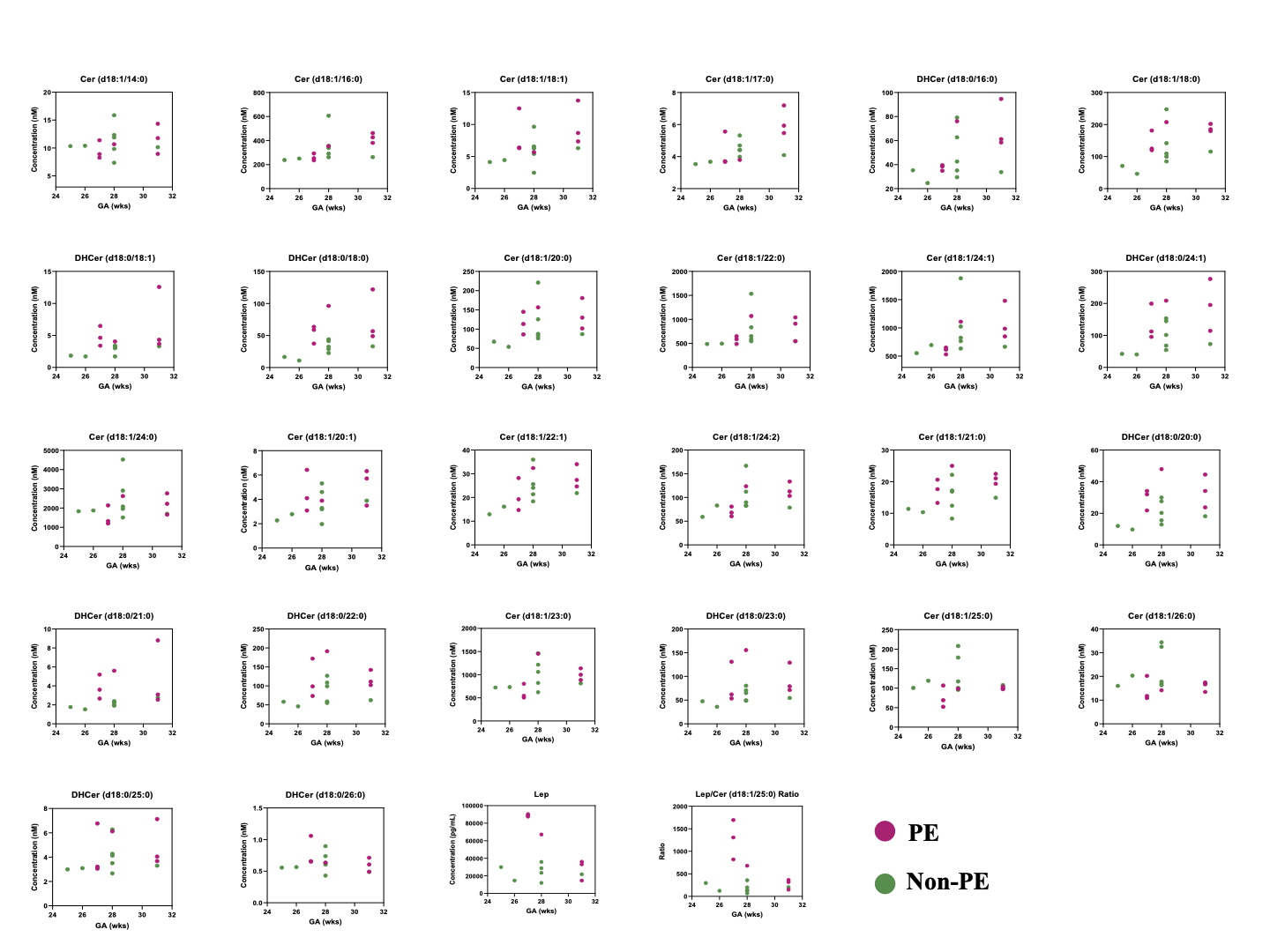

**Appendix 3.** Cohort 1: These scatterplots show maternal serum concentrations of each analyte in preeclamptic (PE) and non-preeclamptic (non-PE) women in the testing cohort. Each dot represents a sample. X-axis represents the time of sample collection. Y-axis represents the serum concentration. Analytes include 24 Cers/DHCers, Lep, and the ratio of Lep/Cer (d18:1/25:0). Lep: leptin. GA: gestational age. Cer: ceramide. DHCer: dihydroceramide.

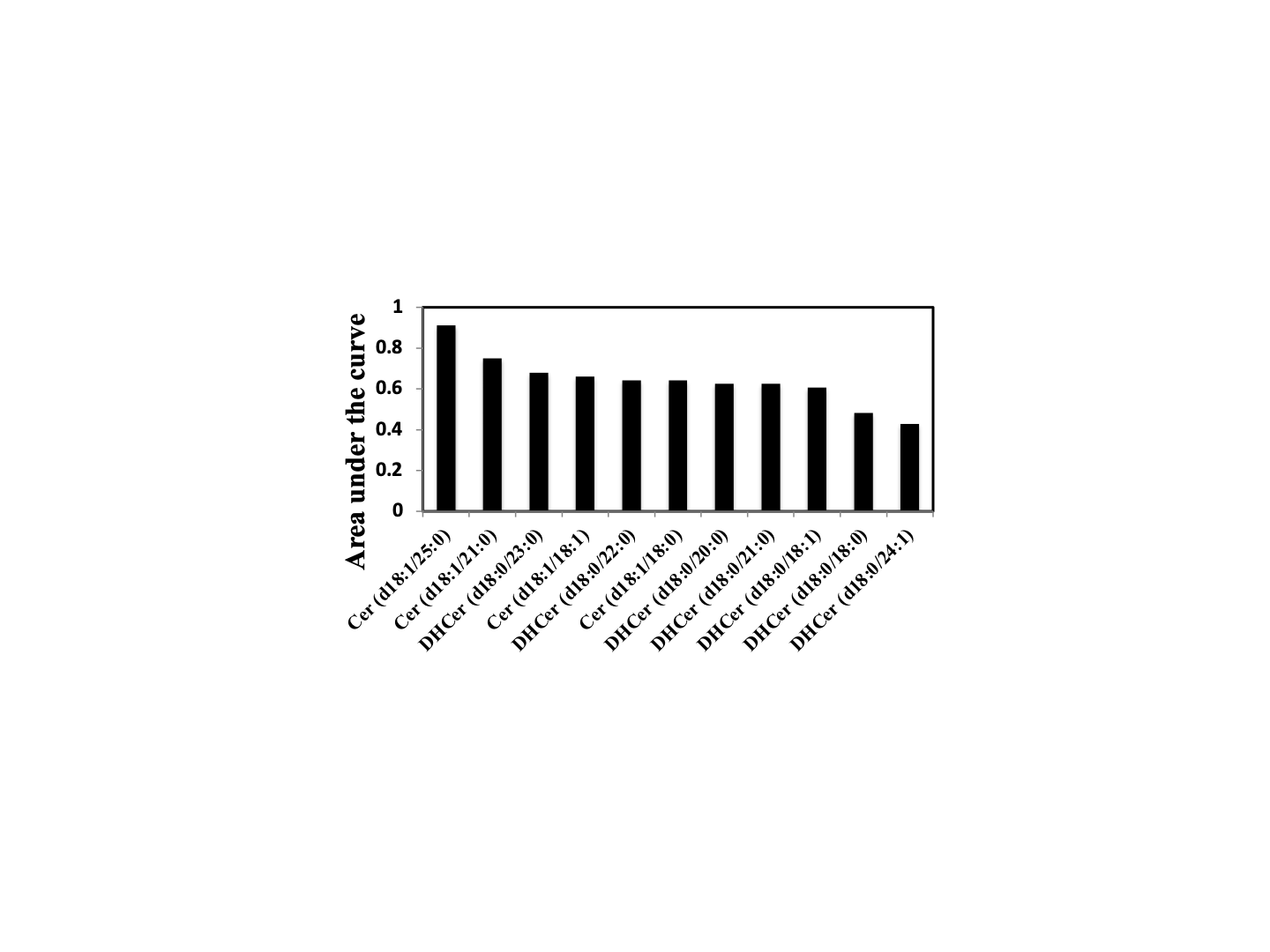

**Appendix 4.** The bar chart shows area under the curves (AUCs) of the ratios of Lep and different Cer/DHCer analytes in classifying preeclamptic (PE) and non-preeclamptic (non-PE) women in the testing cohort. Lep: leptin. Cer: ceramide. DHCer: dihydroceramide.

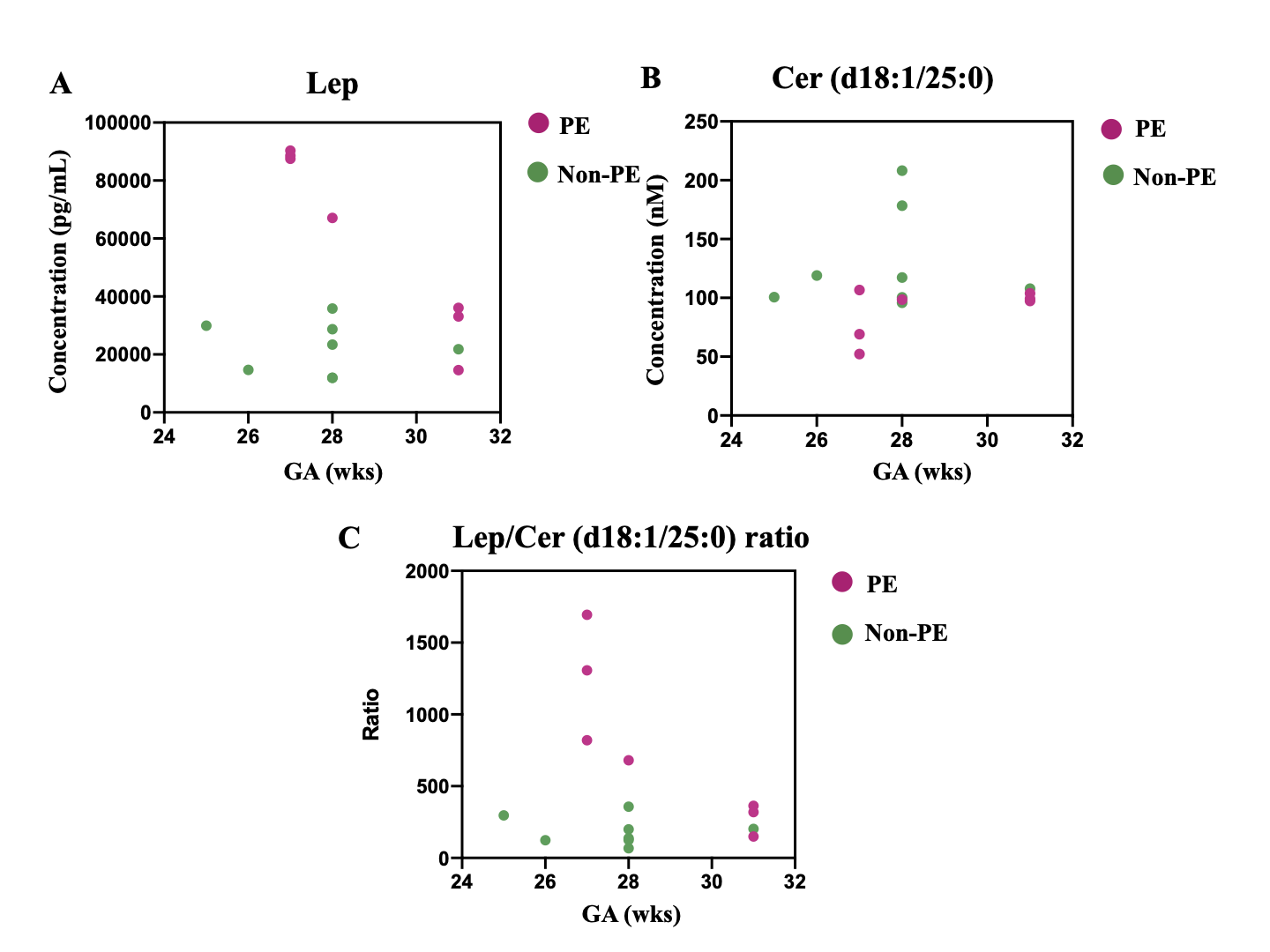

**Appendix 5.** The scatterplots show maternal serum concentrations of (A) Lep, (B) Cer (d18:1/25:0), and (C) Lep/Cer (d18:1/25:0) ratio in the testing cohort. Each dot represents a sample. X-axis represents the time of sample collection. Y-axis represents the serum concentration. Lep: leptin. GA: gestational age. PE: preeclampsia. Cer: ceramide.

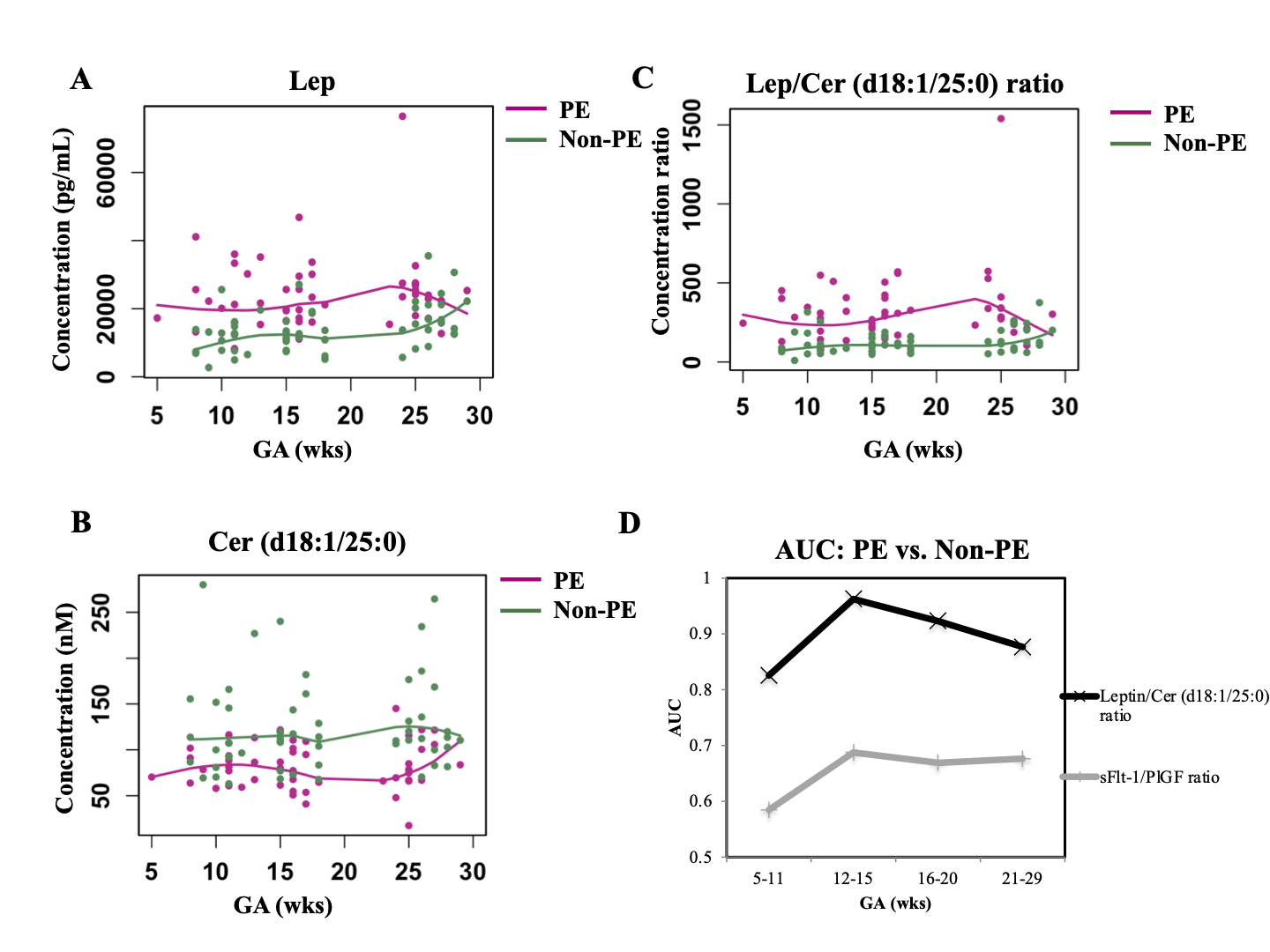

**Appendix 6.** The scatterplots show maternal serum concentrations of (A) Lep, (B) Cer (d18:1/25:0), and (C) Lep/Cer (d18:1/25:0) ratio in the validation cohort, and (D) area under the curve (AUC) values of Lep/Cer (d18:1/25:0) and sFLT-1/PlGF ratios at different windows of gestation. In (A)-(C) each dot represents a sample; X-axis represents the time of sample collection; and Y-axis represents the serum concentration. Loess smooth function was applied. Lep: leptin. GA: gestational age. PE: preeclampsia. Cer: ceramide.

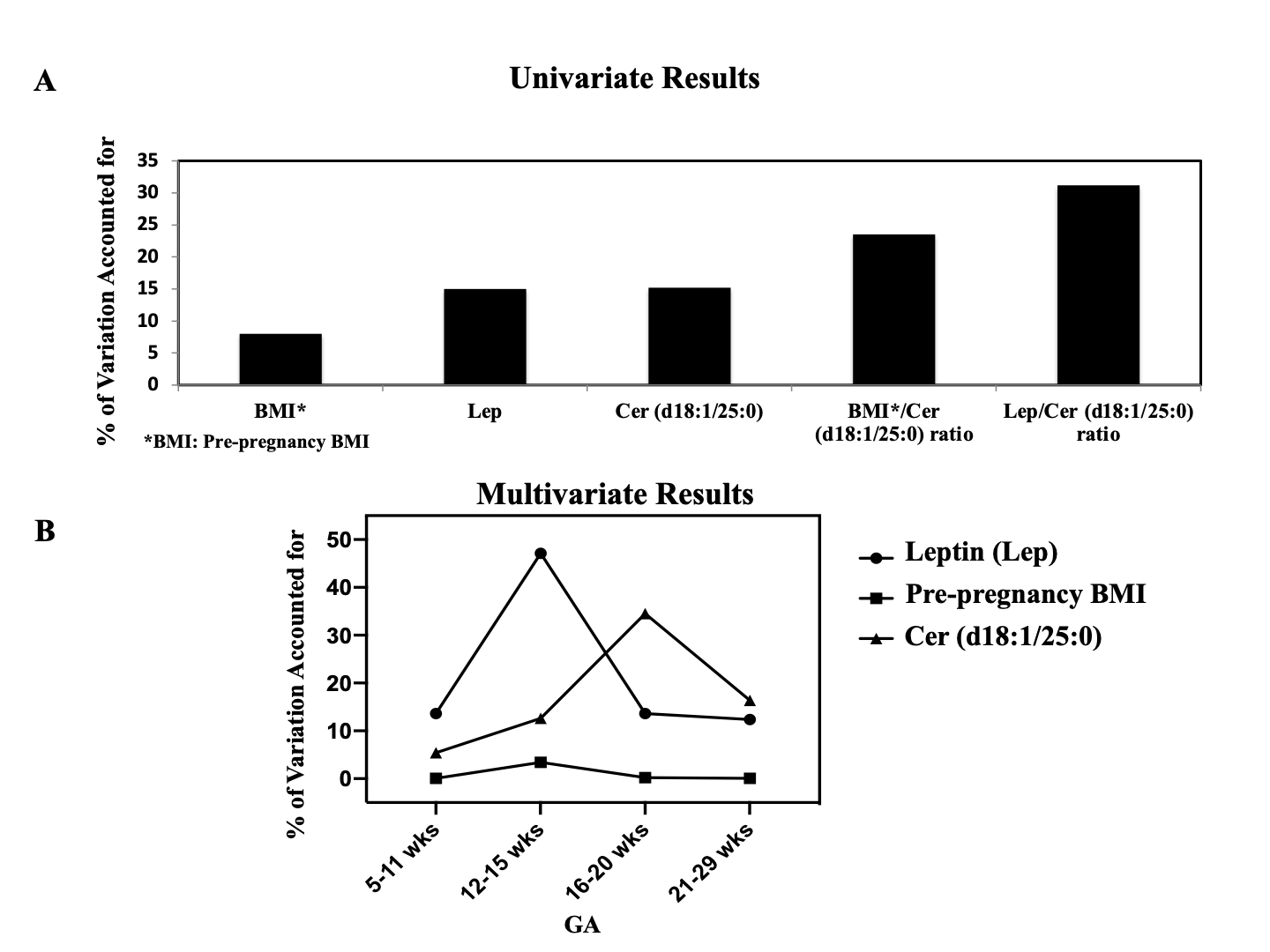

**Appendix 7.** Results of univariable and multivariable analyses on non-obese women (body mass index [BMI] <30 kg/m^2^) in the validation cohort. BMI was measured prior to pregnancy. wks: weeks. GA: gestational age.
